# Clinical validation of Whole Genome Sequencing for cancer diagnostics

**DOI:** 10.1101/2020.10.29.20222091

**Authors:** Paul Roepman, Ewart de Bruijn, Stef van Lieshout, Lieke Schoenmaker, Mirjam C Boelens, Hendrikus J Dubbink, Willemina RR Geurts-Giele, Floris H Groenendijk, Manon MH Huibers, Mariëtte EG Kranendonk, Margaretha GM Roemer, Kris G Samsom, Marloes Steehouwer, Wendy WJ de Leng, Alexander Hoischen, Bauke Ylstra, Kim Monkhorst, Jacobus JM van der Hoeven, Edwin Cuppen

## Abstract

Whole genome sequencing (WGS) using fresh frozen tissue and matched blood samples from cancer patients is becoming in reach as the most complete genetic tumor test. With a trend towards the availability of small biopsies and the need to screen an increasing number of (complex) biomarkers, the use of a single all-inclusive test is preferred over multiple consecutive assays. To meet high-quality diagnostics standards, we optimized and clinically validated WGS sample and data processing procedures resulting in a technical success rate of 95.6% for fresh-frozen samples with sufficient (≥20%) tumor content.

Independent validation of identified biomarkers against commonly used diagnostic assays showed a high sensitivity (recall) (98.5%) and precision (positive predictive value) (97.8%) for detection of somatic SNV and indels (across 22 genes), and high concordance for detection of gene amplification (97.0%, *EGRF* and *MET*) as well as somatic complete loss (100%, *CDKN2A*/p16). Gene fusion analysis showed a concordance of 91.3% between DNA-based WGS and an orthogonal RNA-based gene fusion assay. Microsatellite (in)stability assessment showed a sensitivity of 100% with a precision of 94%, and virus detection (HPV) an accuracy of 100% compared to standard testing.

In conclusion, whole genome sequencing has a >95% sensitivity and precision compared to routinely used DNA techniques in diagnostics and all relevant mutation types can be detected reliably in a single assay.

## Introduction

Needs and complexity in molecular cancer diagnostics are rapidly increasing, driven by a growing number of targeted drugs and developments towards more personalized treatments ^1,2^. Simultaneously, advances in next-generation DNA sequencing technology have greatly enhanced the capability of cancer genome analyses, thereby rapidly progressing diagnostic approaches from small targeted panels to large panels and exome sequencing. Currently, whole genome sequencing (WGS) using tissue and matched blood samples from patients with (metastatic) cancer ^3^ is getting in reach as the most complete genetic tumor diagnostics test. In the context of the Dutch national CPCT-02 clinical study (NCT01855477) Hartwig Medical Foundation has established a national WGS facility including robust sampling procedure and logistics in more than 45 (of the 87) hospitals located across the Netherlands for the centralized analysis of tumor biopsies by WGS. Since the start in 2016, more than 5,000 tumors and matched control samples have been analyzed by WGS, of which the first cohort of 2500 patients has been extensively characterized and described ^4^. Originally, this clinical study aimed to analyse data for biomarker discovery, but with growing clinical demands for more extensive and broader DNA analysis for patient stratification towards targeted treatments ^5^, the scope of WGS is now entering routine diagnostic usage. As part of this development, the required amount of tumor tissue for as well as the turn-around-time of the WGS procedure was decreased, together with implementation of more extensive quality control metrics and independent validation required for accreditation. Currently, there is an ongoing trend towards the availability of only small biopsies, especially for advanced stage cancer where metastatic lesions are sampled using core needle biopsies, with at the same time a growing need to screen for an increasing number of (complex) biomarkers. For future-proof and efficient molecular diagnostics, the use of a single all-inclusive test is preferred over multiple consecutive assays that, together, often take more time, require more tissue and provide a far less complete profile of the molecular characteristics.

To meet the high-quality diagnostics standards, we have optimized and clinically validated the performance of the WGS workflow on fresh-frozen tumor samples, both technically as well as bioinformatically, as these are highly interconnected in determining the precision (positive predictive value) and sensitivity (recall) of the test. The validation efforts include current *standard-of-care* biomarkers (oncogenic hotspots, inactivating mutations in tumor suppressor genes), but also broader analyses of gene fusions and other genomic rearrangements as well as emerging genome-wide or complex biomarkers like tumor mutational burden estimation, microsatellite instability (MSI) ^6^, and homologous repair deficiency (HRD) signatures ^7,8^. Importantly, an *open-source* and data-driven filtering and reporting strategy has been put into place to reduce the wealth of information into a diagnostically manageable size and to provide an overview of all clinically relevant DNA aberrations.

Here we show that WGS has an overall >95% sensitivity (recall) and precision (positive predictive value) as compared to other routinely used tests and that all relevant mutation types can be readily and reliably detected in a single assay. Although WGS required minimal quantity of input material and can be applied pan-cancer, the tumor purity was a limiting factor (requiring >20% tumor cells) as well as the availability of fresh frozen tumor material, that were prerequisites for high-quality results as described here. Together, WGS has now matured from a research technology into an ISO accredited test that is ready to be used for clinical decision making.

## Methods

### Tumor samples

For this study, samples were used from patients that were included as part of the CPCT-02 (NCT01855477), DRUP (NCT02925234) and WIDE (NL68609.031.18) clinical studies, which were approved by the medical ethical committees (METC) of the University Medical Center Utrecht and the Netherlands Cancer Institute. All patients have consented to the reuse of their pseudonymized data for research aimed at improving cancer care.

### Whole Genome Sequencing

Whole Genome Sequencing (WGS) was performed under ISO-17025 accreditation at the Hartwig Medical Foundation laboratory (Amsterdam, the Netherlands). The WGS test used DNA extracted from fresh-frozen or frozen archived tumor tissue (primary or metastatic) and from matching blood samples (reference). DNA extraction is performed on the QiaSymphony (Qiagen, Hilden, Germany) following standard reagents and protocols: 1 ml of blood was used for DNA isolation using the QIAsymphony DSP DNA Midi kit (Qiagen). The QIAsymphony DSP DNA Mini kit (Qiagen) was used for tissue DNA isolation. Next, 50-200 ng DNA was fragmented by sonication on the Covaris LE220 Focused ultrasonicator (Covaris, Brighton, UK) (median fragment size 450 bp) for TruSeq Nano DNA Library (Illumina, San Diego, CA, USA) preparation including PCR amplification (8 cycles). All procedures were automated on the Beckman Coulter Biomek 4000 and Biomek i7 liquid handling robots (Beckman Coulter, Brea, CA, USA). The Illumina HiSeqX and NovaSeq6000 platforms were used for sequencing tumor (∼90x) and blood (∼30x) genomes. No minimal threshold was applied regarding the mean coverage but instead the Gbase sequencing output for the tumor and blood samples had to be >300 and >100 Gb respectively, to be eligible for downstream diagnostic analysis. Additional data quality criteria were: read mapping percentage >95%, reference genome-wide coverage 10x >90% and 20x >70%, and tumor genome-wide 30x coverage >80% and 60x >65%.

### Tumor purity

WGS analysis required tumor samples with sufficient tumor cell percentage (≥20%). Prescreening of eligible tumor samples was performed by manual pathological scoring (pTCP) of Haematoxylin and Eosin stained sections, cut from the same frozen biopsy (following standard formalin-fixed paraffin-embedded (FFPE) protocol) that was used for DNA isolation (to minimize the potential effect of tumor heterogeneity). In addition, a molecular based tumor purity (mTCP) was determined based on the WGS data (see bioinformatics) for optimal analysis and interpretation of the DNA results. The mTCP was also determined after shallow whole-genome sequencing (8-15x coverage depth) to be able to identify tumors with a potential discrepancy in pTCP and mTCP before continuing with “deep” sequencing (∼90-110x). This also allowed prescreening tumors for which no (reliable) pathological assessment was available. Only cases with an mTCP of 20% or more were considered eligible for diagnostics analysis.

### Bioinformatics

Sequencing data was analyzed with an in-house developed open source software-based pipeline. Reliable variant calling by sequencing techniques (especially WGS) depends on a complex, often Bayesian, approach including read quality, variant allele frequency, sequence depth and tumor purity and ploidy. A schematic overview of all the used tools is provided in **Suppl Figure 1**. Sequencing read alignment of matching tumor and blood reference samples was performed using the Burrows-Wheeler Aligner (BWA version 0.7.17). Somatic variant calling (single nucleotide variants (SNV), multi-nucleotide variants (MNV) and insertions and deletions (indels)) between the tumor reference pair was performed using STRELKA (version 1.0.14) with which indels up to 50 bp could reliably be identified ^9^. Larger insertions and deletions (50 bp or more) were detected using the tool GRIDSS (version 2.8.3) as being structural variants. GRIDSS is a structural variant detection tool including a genome-wide break-end assembler and a somatic structural variation caller, and is able to detect genomic break-junctions ^10^.

Variant and gene ploidy aspects were assessed using the AMBER tool (version 3.3) that determined allele copy numbers of heterozygous germline variants in the tumor samples. In combination with COBALT (version 1.7), which determined read depth ratios and copy numbers of the supplied tumor and reference data, information was gathered concerning the local copy number and ploidy for bins of ∼1kb across the tumor genome. In addition, a gender check was performed using the COBALT output based on the observed sex chromosome pattern.

Output from the AMBER (bi-allele frequencies), COBALT (read depth ratios), STREKLA (somatic variants) and GRIDSS (structural variants) was combined in the tool PURPLE (version 2.43) (**Suppl Figure 1)** that was designed specifically for WGS data. PURPLE was able to estimate the purity (mTCP) and copy number profile of a tumor sample by searching for the best genome-wide purity/ploidy fit with the input data. The tool provided tumor purity corrected variant allele frequencies (VAF) and allele specific copy numbers that could be used for detection of loss-of-heterozygosity (LoH) ^11^. Importantly, tumor purity correction allowed for reliable identification of somatic complete loss of a gene (e.g. LOH of *BRCA1* and deep (bi-allelic) deletions of *CDKN2A*). Downstream interpretation of structural variants and the calling and annotation of gene fusions was performed using LINX (version 1.7). This tool was able to group together the individual structural variant calls into distinct events, prediced the local structure of the derivative chromosome and properly classified and annotated events for their functional impact ^11^.

Genome-wide mutational characteristics were determined including the tumor’s mutational load (ML, defined as the total number of somatic missense variants across the whole genome of the tumor) and mutational burden (TMB, defined as the number of all somatic variants per genome Mb). Microsatellite instability (MSI) was assessed using the method described by the MSISeq tool ^6^. In brief, the number of indels was calculated per million bases and occurring in homopolymers of 5 or more bases or dinucleotide, trinucleotide and tetranucleotide sequences of repeat count 4 or more. Samples with an score greater than 4 were classified as MSI.

Homologous Recombination DNA repair-deficiency (HRD) was assessed using the previously described CHORD tool (version 60.02_1.03) ^8^. The CHORD tool is random forest classifier of HRD and was able to distinguish between BRCA1/2-type HRD phenotypes. The main discriminants for HRD were the numbers of deletes with micro-homology and the number of large duplications with length between 1kb and 100kb. CHORD achieved a maximum F1-score (∼0.88) for predicting HRD with a cutoff of 0.5 and samples above this cutoff were classified as HR-deficient ^8^.

Furthermore, the presence of viral DNA was detected using VIRUSBreakend (GRIDSS subtool) that identified viral integrations anywhere in the host genome using a single breakend-based strategy followed by taxonomic classification of the detected viral DNA ^12^.

All code and scripts used for analysis of the WGS data are open source and available at GitHub (https://github.com/hartwigmedical/). The raw and analyzed WGS data used in this manuscript are available for validation and cancer research purposes through a standardized controlled data access procedure (see https://www.hartwigmedicalfoundation.nl/applying-for-data/ for details).

### Orthogonal validation experiments

Independent validation was performed for all to-be-reported types of clinically relevant DNA aberrations, including mutations (SNV, MNV and indels) with specific focus on *BRAF*, gene amplification (*ERBB2* and *MET* as examples) and complete loss of genes (*CDKN2A* and *BRCA1, BRCA2*), microsatellite (in)stability, gene fusions, and viral infection (Human Papillomavirus (HPV) as example). WGS results were retrospectively compared against (as far as possible) routine diagnostic assays performed independently in ISO15189 accredited pathology laboratories. If a clinical assay was not available for the validation purpose, a custom research-use-only test was performed. The following independently performed validation experiments were performed. An overview of the used tumor samples and tumor types for each validation experiment is available as **Suppl Table 1**.

### Validation of SNV, MNV and indel detection

A custom designed (research-use-only) single molecule Molecular Inversion Probe (smMIP) sequencing panel was designed for independent confirmation of variants detected by WGS. The smMIP panel sequencing was designed and processed similar to previous reports (Radboudumc) ^13,14^. In total 415 smMIPs (covering 1.4 kbp) were designed to test 192 randomly selected variants (165 SNVs and 27 indels) that were detected by WGS across 29 tumor samples. smMIP validation was performed using the same isolated DNA as was used for WGS, and analysed by JSI SeqPilot (version 5.1.0) (JSI medical systems, Ettenheim, Germany).

Orthogonal clinical validation of variant detection was performed using 48 samples and compared against a custom-made Oncomine NGS gene-panel (Thermo Scientific), processed independently (double blind) in a routine pathology laboratory under ISO15189 accreditation (Erasmus MC) ^15^. The custom Oncomine assay covered 25.2 kb exonic regions across 40 genes (design (v5.1) available in supplementary data of ref. 15) and was performed using the same isolated DNA as was used for WGS, thereby ruling out potential tumor heterogeneity. Analysis was done using JSI SeqPilot version 5.2.0 and a formal clinical report was generated. Additionally, for 10 samples a comparison was made between the WGS based mutational load (ML) assessment and the Oncomine Tumor Mutational Load (TML) assay (Thermo Scientific, Waltham, MA, USA).

### Validation of copy number assessment

WGS based copy number assessment was validated against fluorescent in situ hybridization (FISH) using COLO829 and a cohort of diagnostic tumor samples. For COLO829, a comparison was made for the ploidy of chromosomes 9, 13, 16, 18, 9p24 (*CD274*/*PDCD1LG2*), and 2q23 (*ALK*) (Amsterdam UMC). Chromosome Enumeration Probes (CEP) for the centromeric region of chromosome 9, 13, 16 and (CEP9, CEP13, CEP16, CEP18) were used, as well as locus specific break-apart probes for 2p23 (*ALK*) fusion (Vysis, Abbott, IL, USA) and 9p24 (*CD274/PDCD1LG2*) fusion (Leica Biosystems, Wetzlar, Germany). Slides were visualized on a Leica DM5500 fluorescence microscope (Leica Biosystems) and for each marker, 100 cells/slide were scored for the percentages of cells with respective numbers of chromosomes (signals) counted.

Diagnostic *ERBB2* copy number readout was validated using 16 tumor samples and using HER2/neu FISH analysis at an independent routine pathology laboratory (University Medical Center Utrecht). Fresh frozen sections for FISH analysis were from the same biopsy used for WGS, or from a matching second biopsy obtained at the same moment. FISH scoring was performed according to guidelines ^16^. For fresh-frozen samples, new sections were fixed using overnight incubation with formalin. Subsequently, routine FFPE FISH protocol was used excluding the xylene deparaffinization step. Slides were used for probe hybridization (LPS001, Cytocell, Cambridge, UK), scanned using the Leica DM6000 scanner and analyzed with Cytovision software (Leica Biosystems). A formal clinical report was generated that was compared with the WGS results, for which the absolute copy numbers detected by WGS were compared with the absolute copy numbers detected by FISH.

In addition to *ERBB2*, WGS-based *MET* copy number readouts were investigated for samples classified as positive for *MET* amplification based on routine chromogenic dual in situ hybridization (DISH) on matching FFPE biopsies. Routine *MET* amplification status was assessed using the *MET* DNP and Chromosome 7 DIG probes (Ventana, Tuscan, AZ, USA) on 5 μm thick sections (SuperFrost slide, Thermo Scientific), according to the manufacturer’s instructions. Samples were classified as positive for MET/CEP7 ratio>2.2.

Detection of complete loss of genes by WGS was validated using *CDKN2A* in which the WGS data was compared against p16 protein expression. CDKN2A/p16 was assessed by IHC on 3 μm thick sections of matching FFPE tumor samples, using the monoclonal primary antibody E6H4 (Ventana).

### Validation of fusion gene detection

Validation of gene fusion detection by WGS was performed against RNA-based Anchored Multiplex PCR NGS assay (Archer FusionPlex Solid Tumor, ArcherDx). Twenty-four samples were selected based on the WGS results to include multiple fusion genes. Matching RNA (200 ng), isolated from the same tissue as the DNA that was used for WGS, was analyzed according to routine pathological procedures (ISO15189 certified) (Erasmus MC). A formal clinical report was generated and compared with the WGS results.

### Validation of microsatellite (in)stability readout

For a set of 50 tumor samples, the microsatellite status was validated using the MSI analysis system (Promega, Madison, WI, USA) and performed at a routine pathology laboratory (Erasmus MC) ^17^ and using the same isolated DNA that was used for WGS. This fluorescent multiplex PCR assays analyzed five nearly monomorphic mononucleotide microsatellite loci (BAT-25, BAT-26, NR-21, NR-24, and MONO-27). Matching tumor and blood samples were analyzed for accurate detection. Both the number of positive loci as well as binary classification of microsatellite instable (MSI) and stable (MSS) were reported. Additional MSI positive cases (n=10) were included in the validation based on routine MMR IHC status (mlh1, pms2, msh2 and msh6) and/or *MLH1* methylation status (MS-MLPA kit, MRC-Holland, Amsterdam, The Netherlands).

### Validation of tumor associated virus detection

WGS based detection of presence of high-risk Human Papillomavirus (HPV) and/or Epstein-Barr virus (EBV) DNA was compared against routine pathological testing (Netherlands Cancer Institute) using the QIAscreen HPV PCR Test (Qiagen) for HPV and EBER IHC for detection of presence of EBV in the tumor (both according to standard protocols). If available, results of routine testing for HPV and/or EBV were used for comparison with WGS. If not available, HPV status was determined retrospectively using an aliquot of the DNA (20 ng) that was used for WGS.

## Results

### Analytical performance

In addition to the orthogonal clinical validation experiments that are described in the next paragraphs, the analytical performance of WGS was continuously monitored using a Genome-in-a-bottle (GIAB) mix-in sample (tumor 30% NA12878: normal 100% NA24385) for which all DNA aberrations were known. The accuracy of GIAB genome-wide variant detection (SNV and short indels) by WGS was very high and stable across different runs and using multiple sequencers (in a time period of eight months) with a precision of 0.998 (range 0.994-0.998) and a sensitivity (recall) of 0.989 (range 0.973-0.990) (**Table 1**). F-scores (combining the precision and recall of the test) for variant detection exceeded the pre-set 0.98 lower limit for high-quality sequencing data (median 0.993, range 0.985-0.994). Direct comparison of all genome-wide somatic base calls (COLO829) between HiSeq and NovaSeq runs indicated a concordant result for 99.99953% of the bases. All discordant bases (1445 out of ∼3.1 billion) were located outside protein coding regions of cancer associated genes (460 genes, 2.33 Mbp) resulting in identical reported results based on both platforms (SNV and indel analysis only). WGS coverage analysis across a set of 25 randomly selected tumor samples indicated stable and high coverage across the entire genome (median coverage after mapping 106x, range 84-130). The protein coding regions of 460 cancer associated genes showed a median coverage of 105x (range 78-134) with 99.68 and 99.29% of all bases covered at least 10x and 30x, respectively (**Table 1**).

**Table 1.**
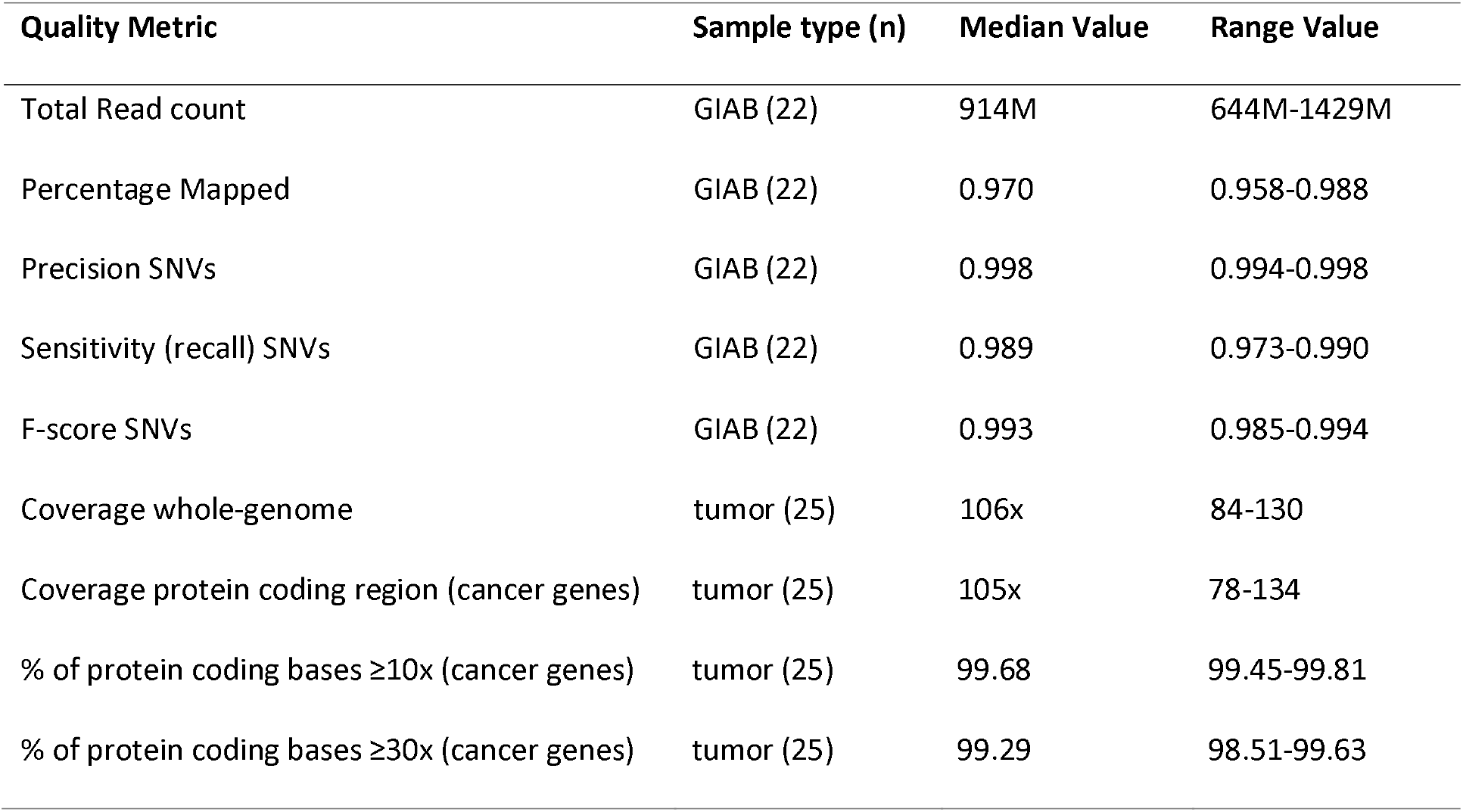
Performance characteristics for clinical-grade WGS using Genome-in-a-bottle (GIAB) and tumor biopsy samples. The GIAB samples (n=21) has been analyzed in duplicate runs using multiple sequencers and across a period of eight months in 2018. Data from 25 randomly selected tumor samples (from 2018) were used for coverage performance. The protein coding region included 460 cancer associated genes (2.33 Mbp in total). The F-score is a measure of a test’s accuracy and is calculated from the precision (true positive / true positive + false positive) and sensitivity (recall, true positive / true positive + false negative) of the test.

To discern the (minimal) required variant read counts (ALT) and variant allele frequencies (VAF, non-purity corrected) of the Bayesian calling pipeline, an analysis was performed to the VAF/ALT of reliable detected non-synonymous variants for 118 cancer associated genes across a set of 2,520 tumor samples ^4^. Out of more than ten-thousand called variants, only 4 variants were based on an ALT count of 4 or less, indicating that for the WGS setup used (combination of wet-lab and bioinformatics) at least 5 ALT reads are required for reliable variant calling, representing a minimal sample VAF of 5% (with a coverage of ∼100x) (**Suppl Figure 2**).

The minimally required tumor cell percentage (purity) for sensitive variant detection was assessed using an in-silico sensitivity model with a pre-set minimal coverage of 100x and ALT read count of 5. Based on this model the minimal tumor purity with a sensitivity >95% to reliably detect a single nucleotide variant was determined as 0.19 (**Suppl Table 2**). This minimally required tumor purity was experimentally confirmed using a dilution experiment (COLO829, performed in duplicate) in which the tumor content was lowered incrementally (mTCP of 100%, 34%, 20% and 13%). All four oncogenic driver mutations that are known to be present in COLO829 (*BRAF* p.Val600Glu, *CDKN2A* p.Gly124fs, *SF3B1* p.Pro718Leu, *TP63* p.Met499Ile) where still all reported for the 20% tumor purity sample, while the 13% sample only showed 3 out of the 4 mutations (missing *CDKN2A* p.Gly124fs).

The reproducibility of the complete workflow was confirmed on two diagnostic cases (non-small cell lung cancer and an undifferentiated pleomorphic sarcoma) in which the replicated tests were started with new library preparations from the isolated blood/biopsy DNA samples and resulted in highly similar molecular profiles with identical diagnostic reports (**Figure 1, Suppl Table 3**).

**Figure 1.**
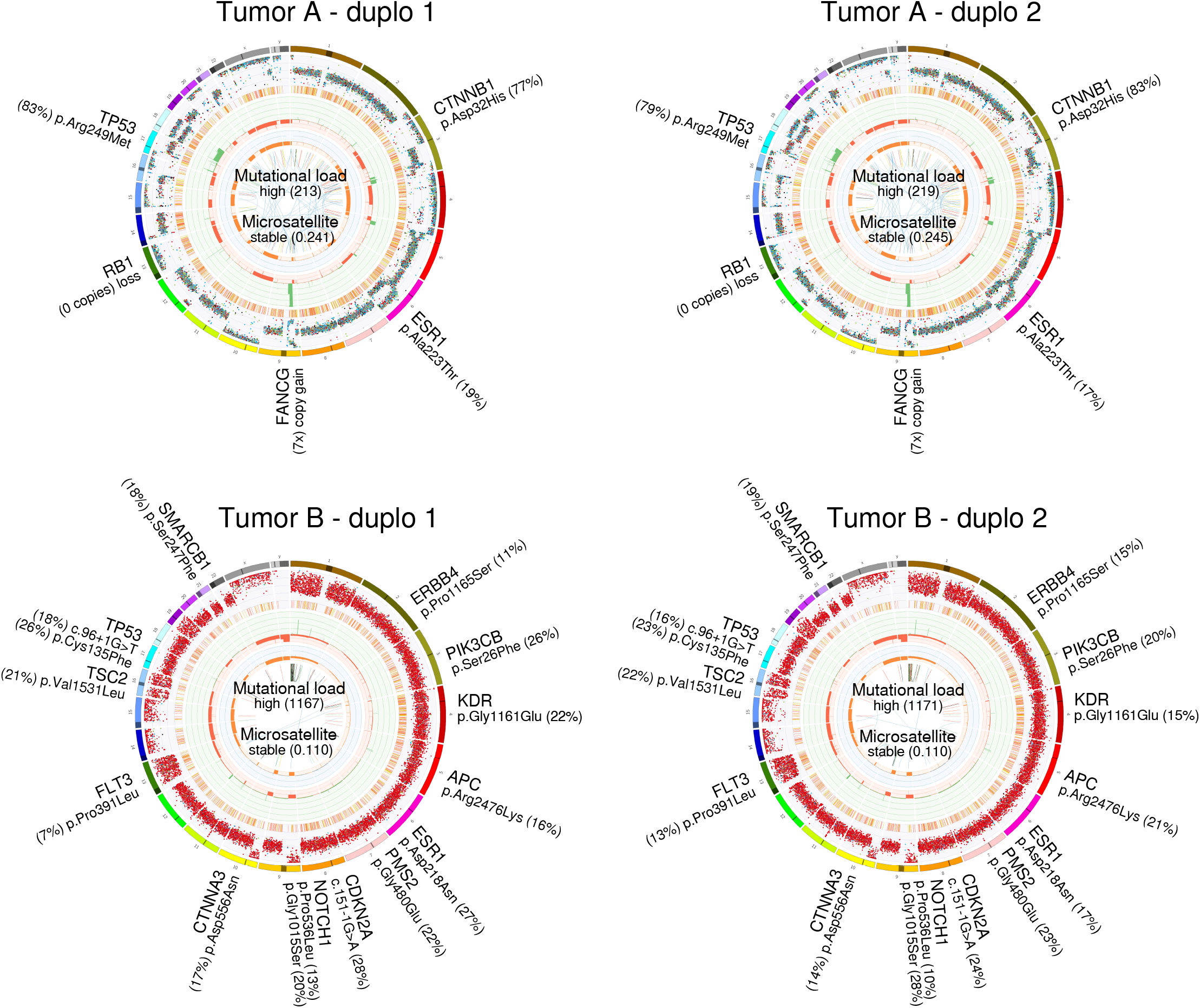
Representation of all tumor specific DNA aberrations as detected using WGS. For each case the complete CIRCOS is shown as well as the reported genomics events, including the mutational burden and microsatellite readout. WGS is performed in duplicate (starting with DNA isolation) for two tumor samples (A, non-small cell lung cancer; B, undifferentiated pleomorphic sarcoma).

### Sample quality, tumor purity and success rate

Samples used for WGS analysis comprised predominantly of freshly frozen fine needle biopsies taken from a metastatic lesion. WGS required at least 50 ng of input DNA and that amount could successfully be isolated from >99% of all eligible biopsies. To determine whether WGS quality is dependent on the (primary) tumor type, a large-scale analysis was performed on the CPCT-02 sample cohort for which samples were collected in 44 different hospitals. Eighty-six percent of all the samples sequenced by WGS (n=2921) passed all quality criteria (n=2520), with a lower success rate for kidney (72.3%), liver (77.3%), and lung (79.1%) cancer patients (**Figure 2A**).

**Figure 2.**
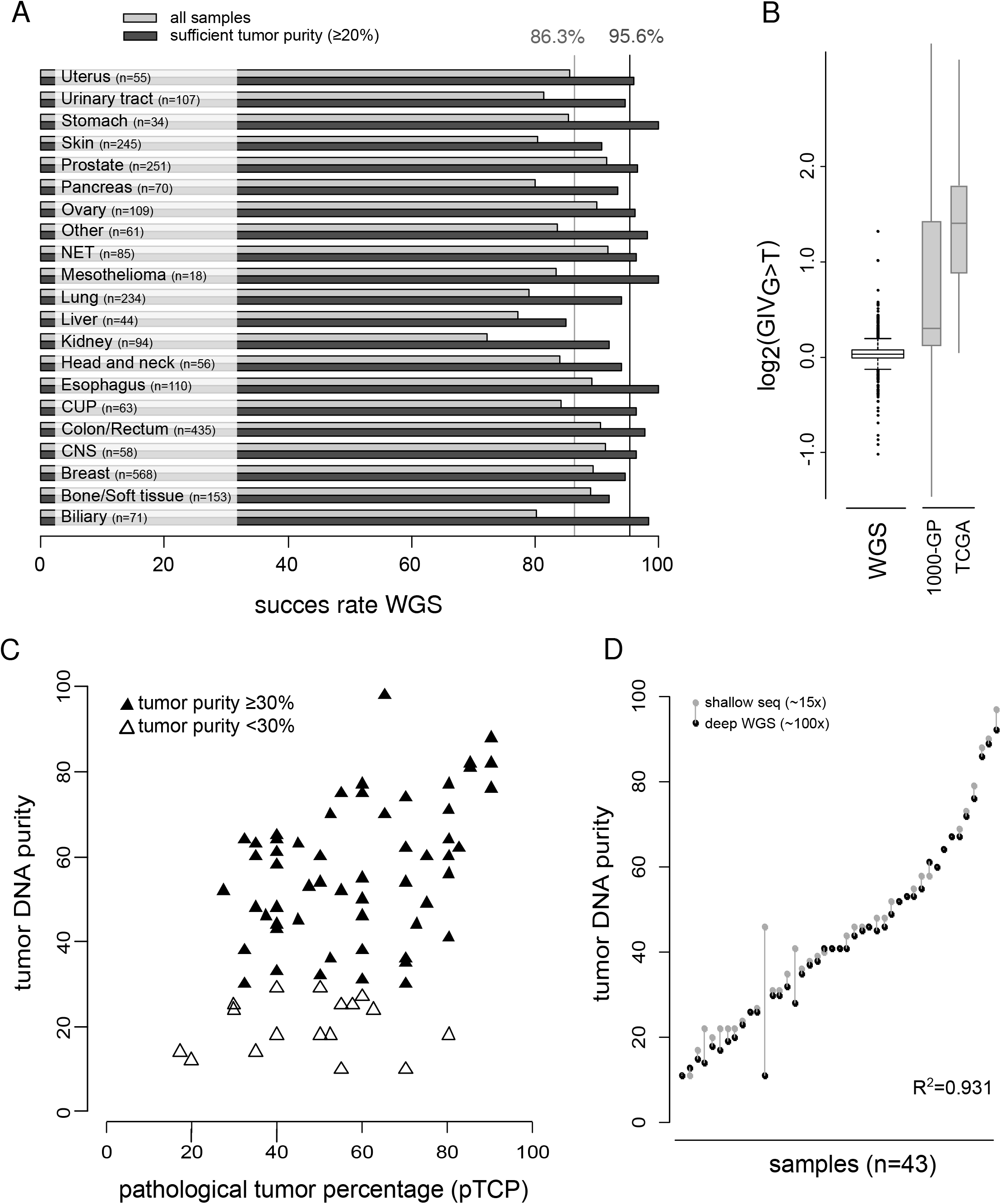
(**A**) WGS success rates for different primary tumor types. Success rates are shown for all samples and for samples that have sufficient tumor content. The average overall success rate across all tumor types is indicated by the vertical lines. (**B**) Global Imbalance Value G to T scores (GIV_G>T_) (n=2520). As a reference the GIV_G>T_ score range is depicted for the 1000 Genomes Project (1000-GP) and a TCGA subset that are described previously ^18^. (**C**) Comparison of pathological tumor percentage scoring (pTCP) with sequencing based tumor DNA purity. (**D**) Comparison of tumor purity assessment using shallow sequencing (grey) (∼15x) and based on deep whole genome sequencing (black) (∼100x) (n=43).

Damaged DNA can cause lower quality sequencing data, as previously described for DNA isolated from FFPE material ^18^. Although damage was expected to be much lower for fresh-frozen samples, the previously described Global Imbalance Value (GIV) score ^19^ was used to directly assess this. The GIV scores are indicative of DNA damage (typically due to oxidation of deoxyguanosine to 7,8-dihydro-8-oxoguanine (8oxodG)) with completely undamaged samples having a GIV_G>T_ score of 1 and severely damaged samples with GIV_G>T_ scores greater than 1.5, resulting in a large excess of false-positive G>T variants due to technical artifacts ^19^. The analyzed set of 2,520 samples showed very low GIV_G>T_ scores with a median of only 1.02 (range 0.495 - 2.495) and only three samples (0.11%) with a GIV score >1.5 (**Figure 2B)**. In comparison, 41% of the 1000 Genomes Project samples showed a GIV_G>T_ score of at least 1.5, while 73% of the TCGA samples (also including FFPE-based samples) showed a GIV_G>T_ score >2 ^19^.

Correct assessment of a sample’s tumor purity is essential for accurate determination of tumor-specific allele frequencies and copy number values. Both manual pathological (pTCP) as well as molecular/DNA-based (mTCP) assessment were performed and using the same fresh-frozen biopsy to minimize potential heterogeneity. The pTCP and mTCP scoring showed a modest but significant correlation for samples with higher tumor content (r=0.40 p=0.002), but this association was absent for samples with lower (<30%) tumor purity (r=0.08, p=0.76) (**Figure 2C**). Additional investigation of sections that were collected before and after cutting the sections (20 tumor samples) revealed intra-tumor heterogeneity but did could not explain all differences (**Suppl Table 4**). Possibly the amount of tumor-infiltrating lymphocytes (each harboring a genome but difficult to quantify in histological slides) plays a role in the observed differences between mTCP and pTCP, especially for tumors with fewer tumor cells.

An insufficient amount of tumor cells was the most prevalent failure rate despite prior pathological prescreening (pTCP>20-30%): 6.4% of samples showed an mTCP between 5-20% and 2.9% showed a seemingly absence of tumor DNA (mTCP <5%). In case reliable pTCP assessment was not available, mTCP calculations based on shallow sequencing data (∼8-15x average coverage) could be used for pre-screening of biopsies eligible for “deep” sequencing. Comparison of mTCPs by shallow and deep WGS (∼90-110x) showed a very good correlation (R^2^ of 0.931, n=43, **Figure 2D**), with an average deviation between both purities of only 3.2% (range 0% to 35% caused by an outlying non-small cell lung cancer case). This result showed that shallow sequencing data was sufficiently reliable for mTCP based estimations and could be used as an alternative for histopathological assessment. When samples were selected with sufficient mTCP (≥20%) and sufficient DNA yield (>50ng), the technical success rate for generating high quality WGS data and reportable outcomes was 95.6%. (**Figure 2A**).

### SNV, indels and mutational burden

Confirmation of variants detected by WGS was initially assessed by a tailored single molecule Molecular Inversion Probe (smMIP) panel sequencing ^13,14^. Across 29 samples, 192 randomly selected variants (165 SNVs and 27 indels, including passenger and driver variants) were sequenced and analyzed by a custom designed smMIP panel (no reliable panel design was possible for 17.6% of the initial selected WGS variants mainly due to the vicinity of intergenic repeat regions). Nearly all (98.4%) of the variants were confirmed by smMIP sequencing indicating a high sensitivity (recall) of the smMIP assay and a high true positive rate of WGS. The observed variant allele frequencies showed a high correlation (R^2^=0.733) between both assays (**Figure 3A**). Three variants could not be confirmed by smMIP: one intergenic SNV (chr3:75887550G>C) due to a double mutation at that position for which the smMIP panel called the other variant (chr3:75887550G>T), and 2 intergenic indels (chr8:106533360_106533361insAC and chr12:125662751_125662752insA).

**Figure 3.**
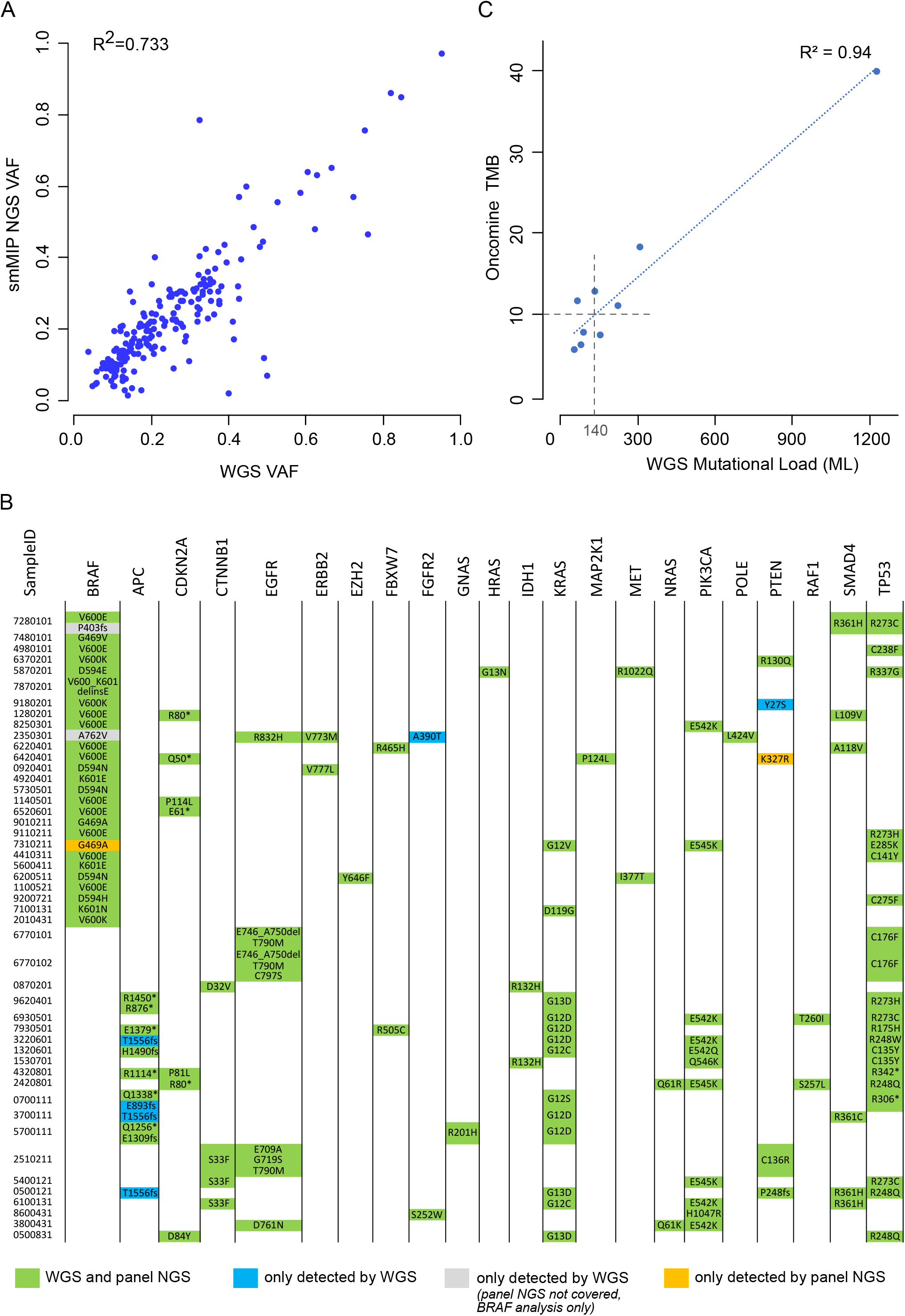
(**A**) Variant allele frequencies (VAF) for SNV, MNV and short indel variants that are detected using WGS and confirmed by smMIP NGS panels sequencing. (**B**) Overview of all protein-changing mutations that are detected by WGS and or the custom-made Oncomine NGS assay. Mutations reported by both assays are marked in green, variants only reported by WGS in blue and only using the panel NGS assay in orange. For *BRAF*, also mutations detected by WGS but which are not included in the panel assay design are shown (in grey). For all other genes, only mutations included in the panel design are considered. (**C**) Comparison of WGS based mutational load (ML) readout with NGS panel based tumor mutational burden (TMB).

Orthogonal clinical validation of mutations in a specific oncogene, *BRAF*, was performed using 48 selected samples and compared against the custom-made Oncomine gene-panel NGS assay (Thermo Scientific). Twenty-five samples showed a *BRAF* exon 15 or exon 11 mutation by WGS that were confirmed by panel NGS (**Figure 3B**). Vice-versa, 26 *BRAF* mutations that were detected using panel-based sequencing were also identified using WGS. A single *BRAF* p.Gly469Ala mutation identified by panel NGS was not confirmed using the WGS analysis due to low mutation frequency (∼2%). WGS identified two less common *BRAF* variants (p.Ala762Val and p.Pro403fs) that were not covered by the used panel design. Both variants were unlikely to result in BRAF activation and were likely passenger variants, especially because both tumors were MSI with a high TMB. All other 20 *BRAF* wild-type samples by WGS were confirmed by panel sequencing.

Next, all somatic non-synonymous mutations across the NGS panel design were evaluated (25.2 kb covering hotspot exons of 40 genes). Combined with the *BRAF* results, in total 138 mutations (121 SNVs and 17 indels) were detected by at least one of the tests of which 136 were reported by WGS and 133 using panel sequencing (**Figure 3B, Suppl Table 5**) resulting in an overall 98.5% sensitivity (recall) and 95.6% precision (positive predictive value) for WGS compared to panel based. A *PTEN* p.Lys327Arg mutation that was identified using the panel, was not reported by the WGS test. Re-analysis of the WGS read data confirmed the presence of this variant with a lower VAF in the tumor (7% with a coverage of 8 out of 116 reads) but also with reduced coverage in the blood reference. This combination affected the Bayesian somatic variant calling algorithm (which depends on information from both the tumor and normal ref samples) and as a consequence no somatic variant could be reliably called. On the contrary, the panel assay did not report a pathogenic *PTEN* variant (p.Tyr27Ser), which was identified by WGS (VAF 12%) using the same input DNA. The variant was present in the NGS panel data (VAF 6%) but was not reported due to incorrect manual curation. The panel did miss identification of the *APC* p.Thr1556fs inactivating mutation in three samples. This *APC* codon lies within a homopolymeric DNA region and the IonTorrent sequencing technology used for the panel sequencing is known to face more difficulties in repetitive DNA regions. Considering the *APC* p.Thr1556fs as true positive results, the WGS precision (positive predictive value) was re-calculated as 97.8%

Although the performance of tumor mutational load (ML) estimations are directly following the performance of accurate non-synonymous variant calling (analytically, ML is only a simple summation of the observed variants), mutational burden readout was compared on 10 additional samples between WGS and Oncomine Tumor Mutational Load (TML) assay (Thermo Scientific). Both readouts showed a high correlation (R^2^=0.94) but this was mainly caused by a single high ML sample (ML > 1200) (**Figure 3C**). Binary classification based on both tests (WGS based ML cutoff of 140 mut vs. TML based TMB cutoff of 10 mut/Mb) indicated a concordance for 7 out of 9 samples (1 sample was not evaluable by Oncomine TML), but also indicated a lower correlation in the cutoff region (R^2^=0.16 when excluding 2 highest ML/TMB samples). This result illustrated the challenge of accurate mutational burden readout using a more limited gene panel as compared to exome or genome-wide measurements, as discussed elsewhere ^20,21^.

Summarized, orthogonal panel NGS validation indicated a high overall sensitivity (recall) (98.5%) and a high precision (positive predictive value) (97.8%) for detection of variants by WGS (SNV (n=121): 98.3% and 98.3%; indels (n=17): 100% and 94.1%, respectively) using fresh-frozen biopsies with ≥20% tumor purity, which was similar as compared to commonly used panel-based approaches on FFPE material ^22^.

### Copy number alterations

WGS chromosomal ploidy and copy number was initially benchmarked against FISH analysis on 6 genomic locations of COLO829 (centromeric region of chromosomes 9, 13, 16, and 18, and 2q23 (*ALK*) and 9p24 (*CD274/PDCD1LG2*)). WGS and FISH analysis showed highly similar purity and ploidy calculations with chr9 showing 4x in ∼55% of cells, chr13 3x in ∼55%, chr18 3x in ∼60%, 2q23 locus 3x in 70-80% and complete diploid chr16 and 9q24 locus for all cells **(Figure 4A**).

**Figure 4.**
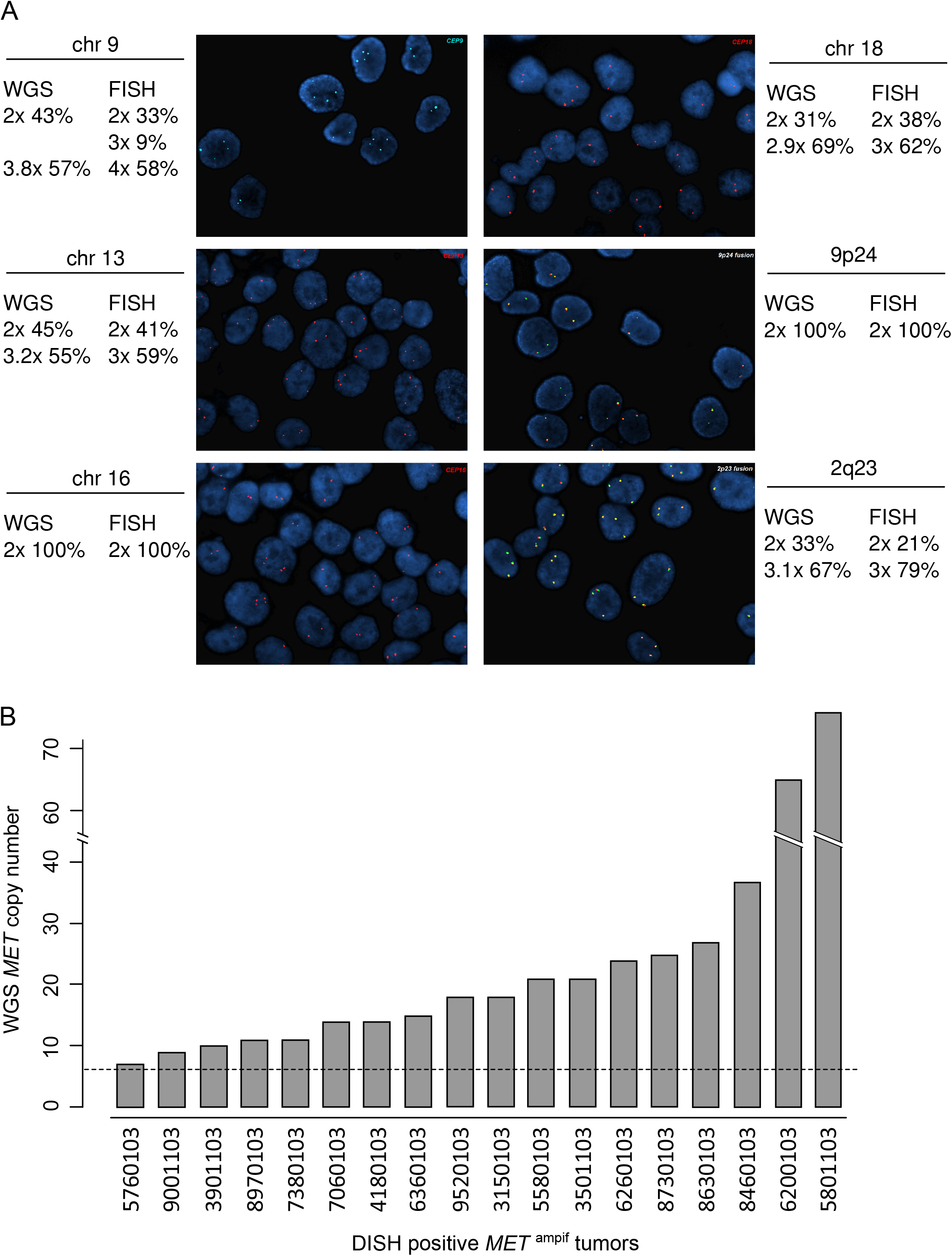
(**A**). Comparison of COLO829 copy number analysis based on WGS and using FISH probes for copy number assessment of chromosomes 9, 13, 16 and 18, and for 9p24 (*CD274/PDCD1LG2*) and 2q23 (*ALK*). For both tests the copy number as well as the percentage of tumor cells is determined. (**B**). WGS based copy number readouts of *MET* of 18 tumor samples that were considered positive for *MET* amplification by routine DISH analysis. The dashed horizontal line represent the 6x copy threshold.

Orthogonal validation of *ERBB2* (*Her2/neu*) amplification detection was performed using 16 samples from various tumor types (**Suppl Table 1**) and including samples with weak and strong amplification levels. WGS *ERBB2* copy gains >6x were considered as actionable amplifications based on previous experience in the CPCT-02 study ^4^ and because this cutoff value is used as eligibility criteria in the Dutch DRUP trail (NCT02925234) for various genes in a pan-tumor setting (e.g *EGFR, ERBB2, MET, FGFR1*) ^23^. Matching fresh frozen sections were analyzed by ERBB2 FISH at an independent routine pathology laboratory (**Table 2**). For one sample (#8700401) FISH analysis failed due to insufficient tumor cells (confirmed by immunohistochemistry), the other FISH results were considered representative. All samples with a WGS copy number greater than 6x were confirmed by FISH to harbor substantial *ERBB2* amplified signals. For copy numbers between 2-6, at best an *ERBB2* gain was observed by FISH but considered insufficient for amplification (classified as *ERBB2* gain or equivocal). A borderline discordant *ERBB2* status was observed for a single case (sample #5550101, FISH 2-4x in 82% of the cells compared to WGS 6x). No technical explanation could be identified, but this might be caused due to tumor heterogeneity between the sections used for WGS and FISH. Of note, this specific case involved a colorectal tumor for which the FISH assay is less common in routine practice.

**Table 2.** *ERBB2* copy number analysis by WGS and FISH. *ERBB2* FISH results were scored solely on tumor cells and categorized as; normal signals, 2-4 signals, 4-6 signals and more than 6 *ERBB2* signals (according to guidelines ^16^). For WGS as well as FISH only absolute copy numbers/counts are used.

| Sample ID | WGS<br>ERBB2 | FISH HER2<br>normal | FISH HER2<br>2-4 | FISH HER2<br>4-6 | FISH HER2<br>>6 | FISH classification |
| --- | --- | --- | --- | --- | --- | --- |
| 3000421 | 9x | 11% | 53% | 7% | 30% | amplification |
| 1810501 | 9x | 8% | 34% | 10% | 48% | amplification |
| 1300611 | 8x | 12% | 31% | 7% | 51% | amplification |
| 8720501 | 8x | 2% | 32% | 15% | 50% | amplification |
| 1210601 | 71x | 2% | 34% | 10% | 54% | amplification |
| 9200111 | 45x | 5% | 10% | 8% | 76% | amplification |
| 3200111 | 43x | 1% | 18% | 4% | 77% | amplification |
| 3300211 | 25x | 6% | 41% | 8% | 45% | amplification |
| 8740201 | 8x | 3% | 22% | 10% | 65% | amplification |
| 5550101 | 6x | 10% | 82% | 7% | 1% | no amplification (equivocal) |
| 3660101 | 5x | 4% | 59% | 37% | 0% | no amplification (gain) |
| 7000711 | 4x | 10% | 20% | 68% | 2% | no amplification (equivocal) |
| 7100331 | 2x | 11% | 63% | 25% | 0% | no amplification (gain) |
| 0200111 | 4x | 28% | 44% | 24% | 4% | no amplification (gain) |
| 5200221 | 6x | 52% | 46% | 3% | 0% | no amplification |

Additional evidence for accurate WGS copy number detection was obtained for *MET*, using 18 samples (**Suppl Table 1**) that had been independently scored as positive for *MET* amplification by DISH analysis during routine diagnostics. All 18 cases showed WGS-based *MET* copy numbers >6 with a large range from 7 to 76 copies and a median 23 copies (**Figure 4B**). Combined, the *ERBB2* and *MET* data showed a high concordance between WGS and ISH analysis (97.0%, 32 of the 33 cases) indicating that WGS reliably detected sufficiently high gene amplifications. For lower gains the concordance showed more variability, but the question remains whether such low gains are biologically and/or clinically relevant ^24^.

To validate the detection of complete bi-allelic loss of genes by WGS, results regarding the presence of complete loss of CDKN2A were compared with routine p16 IHC data of 39 samples. Twenty-two samples with no (zero) intact copies of CDKN2A in the tumor cells as detected by WGS (corrected for tumor purity) were all confirmed negative for p16 expression by IHC (100%, 22 of 22. **Suppl Table 6**). The 17 samples with presence of wildtype *CDKNA2* according to WGS (at least 1 intact wildtype allel) were all found to be positive for p16 IHC. Furthermore, for samples in the 2520 tumor cohort that showed complete loss of all *BRCA1* or *BRCA2* alleles according to WGS, a characteristic HRD profile ^8^ was present in all cases (16 of 16) (**Table 3**), thereby confirming complete bi-allelic *BRCA* inactivation. Of note, this type of bi-allelic *BRCA* inactivation due to complete deletion is challenging to detected reliably by panel NGS, as the PCR amplicon libraries are in such cases based on wildtype BRCA alleles from the normal cells in the tissue samples.

**Table 3.** Homologues recombination deficient (HRD) using the CHORD signature for 16 tumors that showed complete loss of the BRCA1 (n=1) or BRCA2 (=15) gene by WGS. A CHORD score >0.50 is indicative for HRD ^8^.

| Sample ID | Tumor type | Gene deletion<br>(0 copies) | CHORD<br>score | CHORD<br>status |
| --- | --- | --- | --- | --- |
| 5000122 | Breast | BRCA2 | 1.000 | HR-deficient |
| 2410601 | Ovary | BRCA1 | 0.992 | HR-deficient |
| 4610801 | Urothelial tract | BRCA2 | 0.920 | HR-deficient |
| 3001101 | Prostate | BRCA2 | 0.998 | HR-deficient |
| 1110601 | Pancreas | BRCA2 | 0.978 | HR-deficient |
| 7300331 | Breast | BRCA2 | 0.902 | HR-deficient |
| 9930101 | Prostate | BRCA2 | 0.940 | HR-deficient |
| 3000712 | Bile duct | BRCA2 | 0.940 | HR-deficient |
| 7900102 | Prostate | BRCA2 | 1.000 | HR-deficient |
| 2040701 | Prostate | BRCA2 | 0.986 | HR-deficient |
| 6620401 | Breast | BRCA2 | 0.956 | HR-deficient |
| 6000921 | Prostate | BRCA2 | 0.912 | HR-deficient |
| 7330701 | Adrenal gland | BRCA2 | 0.880 | HR-deficient |
| 0100711 | Bile duct | BRCA2 | 0.750 | HR-deficient |
| 1120701 | Prostate | BRCA2 | 1.000 | HR-deficient |
| 3820401 | Prostate | BRCA2 | 0.988 | HR-deficient |

### Fusion genes

Detection of gene fusions by WGS was compared with results obtained with an RNA-based Anchored Multiplex PCR NGS assay (ArcherDx) and was performed independently on 24 samples using matching DNA and RNA from the same biopsy. Samples were selected based on the WGS results to include one or more clinically relevant fusion genes. The Archer NGS assay confirmed the WGS findings for 21 of the 23 samples (91.3%), including fusion of *ALK, NRG1* and *ROS1* (**Table 4**). For one sample no comparison could be made, as the *TMPRSS2-ERG* fusion was not covered by the used Archer FusionPlex assay.

**Table 4.** Fusion genes detected by WGS and the Archer FusionPlex on matching DNA and RNA.

| Fusion gene details | WGS | Archer NGS | Nr | Sample ID |
| --- | --- | --- | --- | --- |
| no fusion | none | none | 7 | 7650101, 1380101, 0590101, 2310011, 9010211, 2510211, 4900321 |
| EIF2AK2 ex12 - ALK ex3 | yes | yes | 1 | 1460201 |
| EML4 ex13 - ALK ex20 | yes | yes | 5 | 4980101, 4980102, 7190101, 5120401, 5120402 |
| EML4 ex2 - ALK ex18 | yes | yes | 1 | 9320501 |
| EML4 ex6 - ALK ex20 | yes | yes | 2 | 6690101, 3430201 |
| SPAG17 ex20 - ALK ex9 | yes | none | 1 | 7330501 |
| EZR ex10 - ROS1 ex34 | yes | yes | 1 | 4500401 |
| GOPC ex8 - ROS1 ex35 | yes | yes | 2 | 2080101, 1410801 |
| MEF2D ex1 - NTRK1 ex2 | none | yes | 1 | 3190101 |
| PTPRF ex11 - NRG1 ex6 | yes | yes | 1 | 1100631 |
| TRPS1 ex1 - NRG1 ex2 | yes | yes | 1 | 4100511 |
| TMPRSS2 ex2 - ERG ex3 | yes | n/a* | 1 | 0530701 |
| Total |  |  | 24 |  |

\* TMPRSS2-ERG fusions are not included in the used Archer FusionPlex assay.

A *NTRK1* fusion detected by Archer NGS (*MEF2D-NTRK1*: (22 reads, 60% VAF) could not be identified using WGS, possibly due to a complex structural variation pattern involving multiple break-junctions in the intronic regions and thus more difficult to call using WGS data compared to analysis of RNA. Vica versa, one fusion (*SPAG17-ALK*) detected by WGS showed no evidence in the tumor RNA. Although based on fusion at DNA level a viable in-frame fusion protein was predicted, it can very well be that the corresponding RNA was expressed at low levels (e.g. due to temporal or spatial expression variation) that are insufficient for reliable detection by the Archer assay.

### Microsatellite instability (MSI)

WGS microsatellite (in)stability classification was validated independently using 60 samples including multiple tumor types (**Suppl Data 1**) and compared to the routinely used 5-marker PCR MSI panel ^17,25^ (50 samples) or compared to MMR/MLH1-methylation analysis (10 samples). Assessment of MSI by WGS was defined as the number of small indels per million bases occurring in ≥5-mer homopolymers and in di-, tri- and tetranucleotide repeats ^6^. The cohort of 2520 tumor samples showed an average MSI score of 1.11 with the vast majority of samples having a low score and a long tail towards higher MSI scores (range 0.004 to 93, **Figure 5A**). 2.7 percent of the samples were classified as MSI using a cutoff of 4 (cutoff was based on the apparent bi-nominal distribution of the MSI scores). On the validation set (n=60) the sensitivity of WGS MSI classification was 100% (95%CI 88.8-100%) with a precision of 94% (95%CI 84.8-93.9%) and a Cohen’s kappa score of 0.933 (95%CI 0-732-0.933). In addition to the binary MSI/MSS concordance, the MSI score correlated with the number of positive PCR markers in which samples with only 1 or 2 positive PCR markers showed a marginal MSI score (**Figure 5B**).

**Figure 5.**
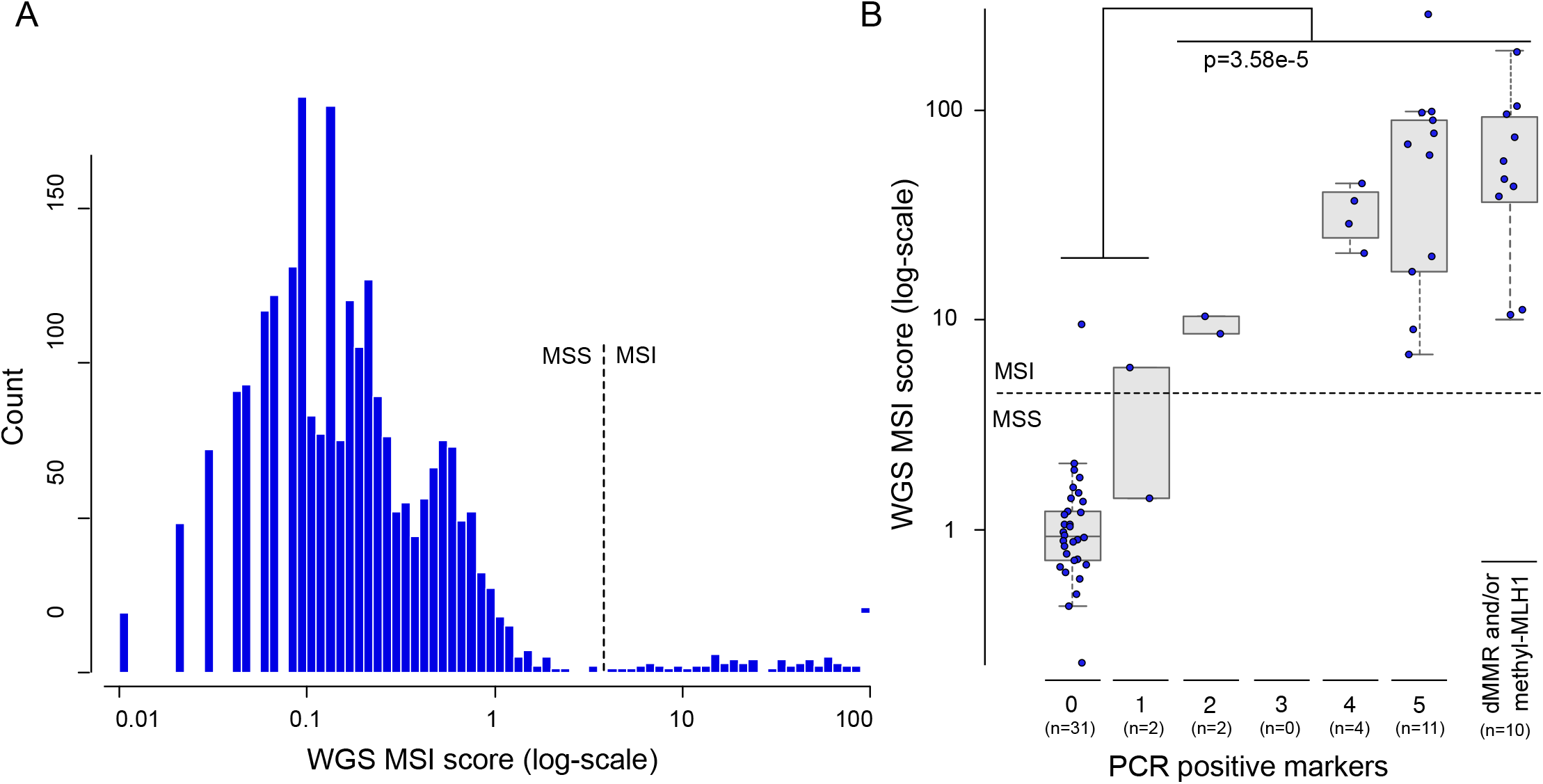
(**A**) WGS based microsatellite instability (MSI) quantification across a cohort of 2520 metastatic cancer samples. (**B**) WGS MSI readout compared to the 5-marker PCR based test using an independent set of 60 validation samples.

One of the two discordant cases was a lymphoma sample (#2300211) with a complex pathology showing 1/5 positive PCR markers (classified as MSS) but a WGS score of 5.9 (classified MSI). IHC analysis showed no substantial loss of MMR proteins although WGS analysis indicated a somatic *PMS2* p.Ile193Met variant in combination with a likely inactivating *PMS2* structural variant. The p.Ile193Met mutation is classified with a high prior in de Leiden Open Variant Database (LOVD, https://databases.lovd.nl/shared/variants/PMS2) and thus likely represents a pathogenic variant. Both the MSI PCR test as well as the MMR IHC had not been validated for use in lymphoma cases so a definitive conclusion remained difficult. The second discordant case (#0740103), a colorectal cancer sample with a WGS MSI score of 9.7 but without a positive PCR marker (0.5 markers) showed a hypermutation phenotype (ML 8050, TMB 601) and harbored two *POLE* mutations (p.Phe1435Val and p.Ser459Phe). Although technically MSS by the routine PCR assay (and thus considered a discordant validation result) the sample was likely a hypermutator with a DNA repair deficiency.

### Tumor-genome viruses

Recently it has been shown that the presence of viruses can be detected with great accuracy using WGS ^26^. Validation of viral detection focused on Human papillomavirus (HPV) due to the prevalence and clinical importance, and the availability of routine testing (e.g. QIAscreen HPV PCR assay, Qiagen). Thirty-seven tumor samples were used for independent validation between WGS and PCR assay including 24 HPV positive and 13 negative (**Suppl Table 1**). WGS HPV status was in concordance with standard pathology assessment for all 37 cases (100% accuracy, 95%CI) with a Cohen’s kappa score of 1.00 (95%CI 0.70-1.00). HPV high-risk types were concordant between both tests for 21 of the 24 positive cases (**Table 5**). For three samples that were classified as ‘high-risk other’ using the PCR assay, WGS analysis indicated either HPV type 16 (#9360103, 3790103) or type 18 (# 6920103). This appeared more a classification than a detection error, but for which the cause remained elusive.

**Table 5.** Detection and typing of HPV in tumor biopsies using WGS and PCR analysis.

| PCR HPV result | WGS HPV result | nr | Sample ID |
| --- | --- | --- | --- |
| no high-risk HPV | no HPV detected | 13 | 4600103, 6600103, 2810103, 0120103, 3120103, 5220103, 2720103, 1920103, 5530103, 6930103, 1570103, 1190103, 7411103 |
| HPV high-risk type 16 | HPV high-risk type 16 | 19 | 2350103, 1360103, 7201103, 9501103, 6601103, 5311103, 3101103, 6960101, 8980101, 7000103, 7100103, 2110103, 7410103, 0710103, 5720103, 4640103, 9950103 |
| HPV high-risk type 18 | HPV high-risk type 18 | 4 | 3990101, 2900103, 0750103, 0701103 |
| HPV high-risk other | HPV high-risk type 16 | 2 | 9360103, 3790103 |
| HPV high-risk other | HPV high-risk type 18 | 1 | 6920103 |
| Total |  | 37 |  |

In addition to the orthogonal HPV validation, six samples, with presence of EBV viral DNA based on WGS analysis (Human gammaherpesvirus 4, NC_007605.1), were assessed by EBER IHC. Interestingly, the three cases with seemingly integrated EBV DNA by WGS scored positive for EBER, while the 3 non-integrated EBV cases were scored as negative for EBER IHC.

## Discussion

During the past few years, whole genome sequencing (WGS) and the associated data analysis and interpretation has matured from a research-use-only tool to a diagnostic-level technology ^27^. Together with the clinical need to screen for an increasing number of (complex) biomarkers in an increased number of tumor types (or even pan-cancer) ^1,23^ and the often limited available biopsy tissue, the use of a single all-inclusive DNA test is a more than welcome development for efficient molecular diagnostics. Here we report on (retrospective) orthogonal validation efforts of WGS on fresh-frozen biopsies and show, to our knowledge for the first time, that the performance of WGS using biopsy with at least 20% tumor cells is equal to the range of routinely used diagnostic tests with technical concordances of >95%. More specifically, we show that a single WGS-based tumor-normal test can provide information regarding: 1) actionable small variants (SNV and indels, routinely detected by targeted panel tests); 2) gene amplifications (FISH); 3) fusion genes (FISH or RNA panels); 4) microsatellite instability (amplicon fragment analysis); 5) viral infections (PCR) and 6) tumor mutational load determination (larger NGS panels). The turn-around-time has been reduced towards a clinically acceptable maximum of 10 working days comprising a minimal net processing time of one day sample registration and DNA isolation, one day library preparation, two days sequencing and two days data analysis and report generation.

Currently, WGS still requires a tumor content that is higher than focused panel based approaches (minimal 20% for WGS versus 5-10% for panel NGS). This limitation is caused due to a lower sequencing depth by WGS and it’s associated costs, but with ongoing developments, it is anticipated that WGS with ∼250x coverage will become feasible for such samples in the next coming years.

The biggest challenge to start using WGS in routine practice is the need of fresh-frozen (or freshly lysed) samples as this will, for most hospitals, require an adaptation in the pathology laboratories that are currently mostly FFPE orientated. The feasibility of implementing WGS in routine practice is currently being evaluated in a prospective clinical validation study ^28^.

The high performance of WGS is primarily the result of two important aspects that are fundamentally different from most current diagnostics procedures for cancer. First, the use of fresh frozen tumor material yields consistent high quality DNA and sequencing results. Second, parallel processing of the patient’s fresh blood sample to serve as a control/baseline for the matching tumor sample. This way, all germline variants can be automatically subtracted and tumor specific changes can be precisely pinpointed. Even across a focused set of ∼500 cancer associated genes ^4^, the bulk of all missense variants observed in the tumor are in fact inherited germline polymorphisms without clinical significance, making comprehensive (manual) tumor-only interpretation and filtering a daunting task. This challenge is not unique for WGS but in principle also applies for (large) NGS panels ^29,30^. Filtering of germline variants using population database information remains challenging due to various reasons (e.g. biases toward to Caucasian population and rare or sub-population specific variants), and the impact on TMB measurements is likely large when germline and somatic variants cannot be discriminated accurately.

Bioinformatics and high-end reporting tools are essential for data-rich assay. Following WGS, the complex whole genome data and results should (again) become manageable and understandable for the end-users (e.g. pathologists, medical oncologists, treating physicians) requiring a delicate balance between what can be detected and what should be reported. To facilitate downstream interpretation, the WGS setup described here ranks (sorts) all observed non-synonymous variants based on calculated oncogenic driver likelihoods ^4^. This strategy allows for a focus on the oncogenic high-driver events, while still providing all information on likely passenger variants (median/low-drivers). For reporting of gene amplification, the tumor’s average ploidy is used as a filter to avoid reporting too many increased copy number events due to whole genome/chromosome duplications. Clinical annotation of the observed DNA aberrations (mutations in high-driver genes, fusions, CNVs) was performed by automatic integration of open-source knowledgebases (CIViC ^31^, OncoKB ^32^ and CGI ^33^) for which only evidence items with convincing clinical relevance (level A+B) were included. Information regarding potential active (and recruiting) clinical trials is integrated in the reporting using a curated national (Dutch) clinical study registry (https://iclusion.com, last accessed 1-feb-2021).

DNA sequencing tests are often performed as laboratory-developed tests (LDTs) and the technical parameters, validation requirements and quality assurance are typically governed by national regulation and legislation that can differ. Various expert groups have drafted guidelines and recommendations for the standardization of multigene panel testing ^2,34-36^ and for our validation efforts we have followed the guidelines for setup and validation of (new) sequencing tests in ISO-accredited pathological laboratories in the Netherlands. However, with the ongoing approval of NGS panel assays by the FDA (https://www.fda.gov/medical-devices/vitro-diagnostics/list-cleared-or-approved-companion-diagnostic-devices-vitro-and-imaging-tools, last accessed 1-feb-2021) and the upcoming new European Regulations for in-vitro diagnostic medical devices IVDR (2017/746) in 2022 ^37^, it is anticipated that (whole) genome sequencing tests will become regulated following international guidelines, standardization and quality schemes. Clinical validation, as described here, by comparison with common standards (despite that no “gold standard” exists) will be a key component of such regulations.

With the increase in (technical) sequencing capabilities, the bioinformatics part (‘dry-lab’) has become essential for a good analysis and interpretation of the sequencing data of WGS but also for the emerging larger comprehensive panels. Traditionally, (hospital) laboratories have focused most on the wet-lab performance and automation but it has become clear that the downstream bioinformatics, and the data infrastructure to handle (and store) all data, are equally important. All the analysis tools and reporting software should also meet the requirements under CE-IVD and ISO regulations and have to be maintained by a dedicated team to ensure diagnostic continuity.

With the rapid development of more targeted drugs and their associated biomarkers, it is next to standardization of the (complex) test results, important to be able to efficiently and quickly add new biomarkers/genes to the clinical reports (e.g. *NRG1* and *NTRK* fusions and *PIK3CA* activating mutations). WGS will allow such a rapid and efficient co-development of (all) future diagnostic DNA markers, because it ‘only’ requires an update of the bioinformatics and reporting aspects, without the need of laborious and costly new test developments or adaptations of panel designs including the required laboratory analytical validation experiments. In addition, the data from previously tested patients can, in principle and upon request from the treating physician, be reanalyzed for the presence of the (all) new biomarkers and recontacting of the patient can be considered ^38^.

In certain (complex) cases, comprehensive analysis of all genomic aberrations at DNA level might still be insufficient to provide a full picture of molecular tumor characteristics. For example, a gene fusion that is considered in-frame and viable for a fusion protein might still not be expressed and could be considered a biological false-positive finding (while technically correct). Also, the presence of integrated viral DNA (measured as viral integration sites) does not always result in active viral gene expression ^39^. In such situations, analysis of the transcriptome using whole transcriptome sequencing in addition to WGS can provide a (more) complete molecular characterization of a tumor.

Comprehensive DNA and/or RNA screening can likely also assist in a (more) detailed classification and diagnosis of tumor types. Currently, tumor classification still relies on histopathological investigation but progress has been made to also start using genomic classifications, especially in the context of rare cancer and cancers with unknown primary (CUP) ^40,41^. We can envision a future in which WGS does not only provide information on possible treatment options but also provides another piece of the puzzle to resolve a complex diagnosis.

Setting aside the direct impact WGS can have for clinical use and comprehensive screening for clinical study eligibility, a whole-genome view of the tumor will yield a wealth of valuable research data and provide the opportunity to increase our insights in oncogenic processes and to better explain or predict the response to targeted or immunotherapy. Such a learning-health-care system, where we learn from today’s patients will greatly enhance our understanding of this complex disease and facilitate the discovery of newly identified (complex) biomarkers, targeted therapies, and improved treatment decision making for future patients.

## Data Availability

The raw and analyzed WGS data used in this manuscript are available for validation and cancer research purposes through a standardized controlled access procedure (see https://www.hartwigmedicalfoundation.nl/applying-for-data/fordetails).

https://www.hartwigmedicalfoundation.nl/applying-for-data/

## Acknowledgements

The authors would like to thank Peggy Atmodimedjo, Isabelle Meijssen, Ronald van Marion and Hanna Schoep for (technical) assistance with collecting the data, Sandra van den Broek, Nina Jacobs and David Koetsier for data analysis support and Immy Riethorst for sample logistics. This publication and the underlying study have been made possible partly on the basis of the data that Hartwig Medical Foundation and the Center of Personalised Cancer Treatment (CPCT) have made available to the study.

