## Supplementary Figure 1 for "Clinical validation of Whole Genome Sequencing for cancer diagnostics"

Schematic overview of the used bioinformatics tools for WGS analysis. All code and scripts used for analysis of the WGS data are available at GitHub:

BWA (version 0.7.17): http://bio-bwa.sourceforge.net/
STRELKA (version 1.0.14): https://github.com/Illumina/strelka
GRIDSS (version 2.8.3): https://github.com/PapenfussLab/gridss
VIRUSBreakend (GRIDSS subtool):
https://github.com/PapenfussLab/gridss/blob/master/VIRUSBreakend_Readme.md
AMBER (version 3.3): https://github.com/hartwigmedical/hmftools/tree/master/amber
COBALT (version 1.7): https://github.com/hartwigmedical/hmftools/tree/master/count-bam-lines
PURPLE (version 2.43): https://github.com/hartwigmedical/hmftools/tree/master/purity-ploidy-estimator
LINX (version 1.7): https://github.com/hartwigmedical/hmftools/tree/master/sv-linx
CHORD (version 60.02_1.03): https://github.com/UMCUGenetics/CHORD

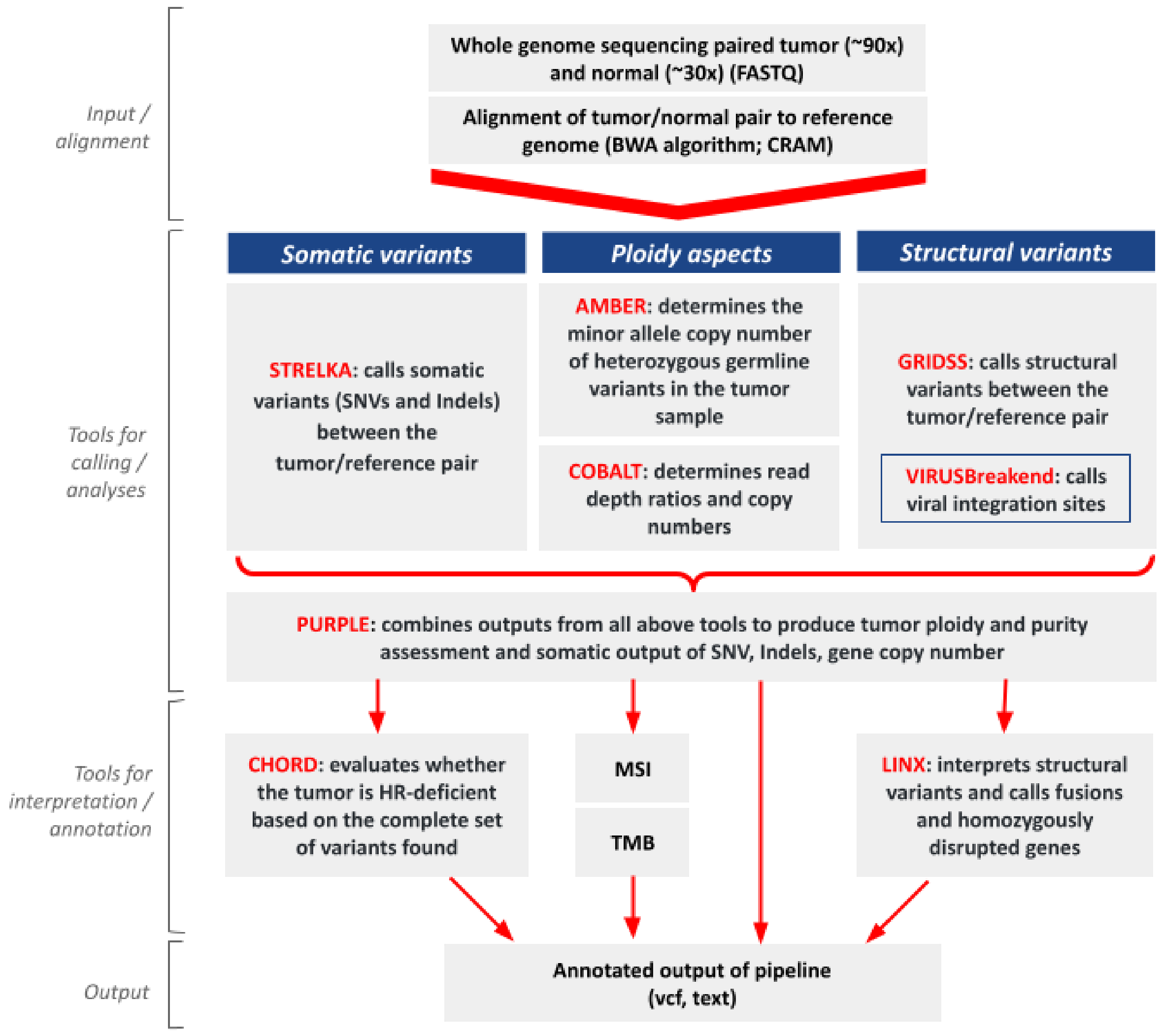
