## Supplementary Figure 2 for "Clinical validation of Whole Genome Sequencing for cancer diagnostics"

Variant allele frequencies (VAF) (not corrected for tumor puirty) and the number of variant (ALT) reads observed across 10,772 reliable detected non-synonymous variants for 118 cancer associated genes across a set of 2,520 tumor samples.

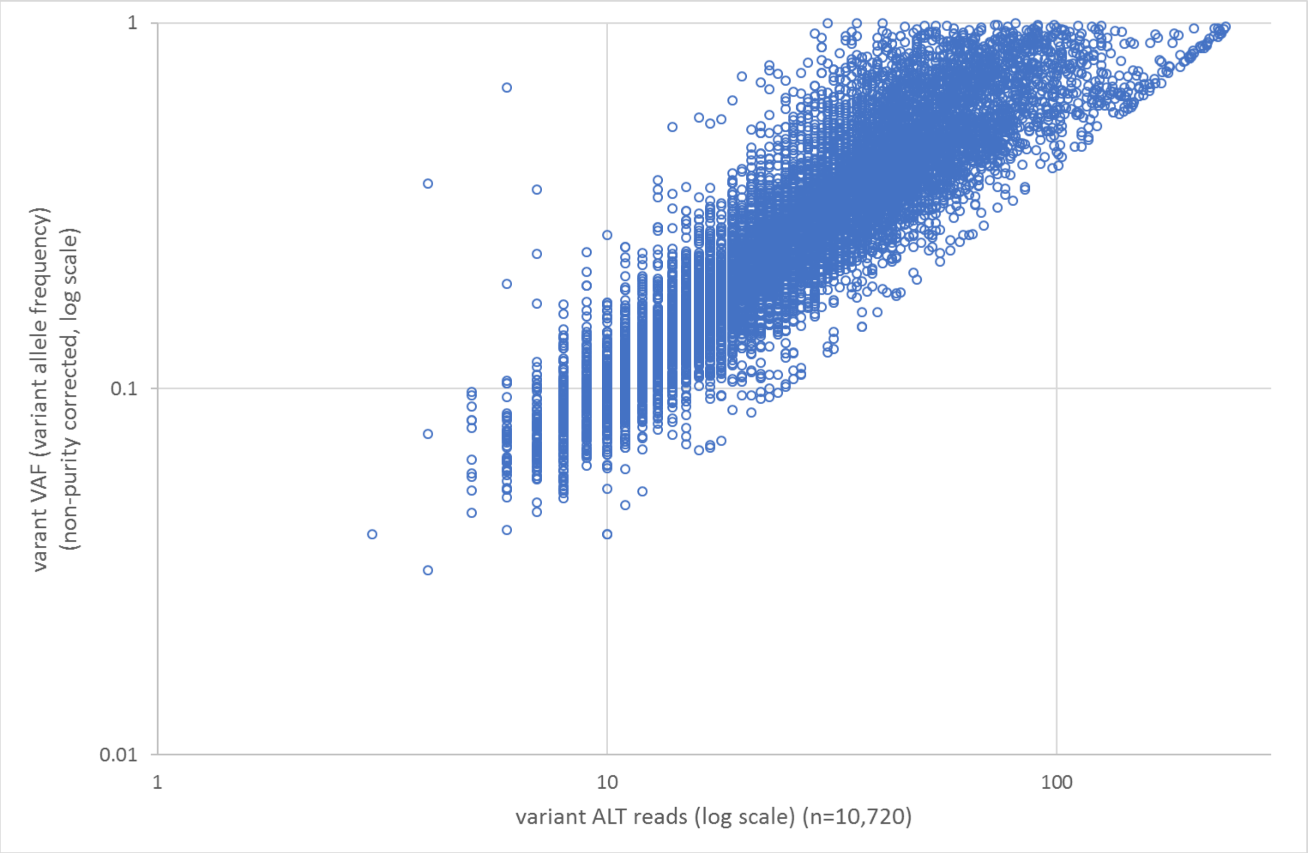
