## Supplementary Table 1 for "Clinical validation of Whole Genome Sequencing for cancer diagnostics"

| SampleID | Tumor type | Validation test | Institute |
|---|---|---|---|
| 0001103 | Bladder cancer | CDKN2A/p16 IHC | Netherlands Cancer Institute |
| 0010701 | Prostate cancer | smMIP panel sequencing (RUO) | Radboudumc |
| 0120103 | Head and Neck cancer | QIAscreen HPV PCR Test | Netherlands Cancer Institute |
| 0180201 | Melanoma | Oncomine TML assay | Erasmus MC |
| 0200111 | Breast cancer | HER2/neu FISH | University Medical Center Utrecht |
| 0210701 | Esophageal cancer | MSI analysis system | Erasmus MC |
| 0301103 | Colorectal Cancer | MMR IHC / MLH1 MS-MLPA | Netherlands Cancer Institute |
| 0410701 | Prostate cancer | MSI analysis system | Erasmus MC |
| 0450103 | Thymoma and Thymic Cancer | CDKN2A/p16 IHC | Netherlands Cancer Institute |
| 0480103 | Lung cancer | CDKN2A/p16 IHC | Netherlands Cancer Institute |
| 0500121 | Colorectal cancer | Oncomine NGS gene-panel (custom) | Erasmus MC |
| 0500831 | Pancreatic cancer | Oncomine NGS gene-panel (custom) | Erasmus MC |
| 0530701 | Prostate cancer | Archer FusionPlex Solid | Erasmus MC |
| 0590101 | Lung cancer | Archer FusionPlex Solid | Erasmus MC |
| 0640103 | Colorectal Cancer | MMR IHC / MLH1 MS-MLPA | Netherlands Cancer Institute |
| 0700111 | Colorectal cancer | Oncomine NGS gene-panel (custom) | Erasmus MC |
| 0701103 | Cervical cancer | CDKN2A/p16 IHC | Netherlands Cancer Institute |
| 0701103 | Cervical cancer | QIAscreen HPV PCR Test | Netherlands Cancer Institute |
| 0710103 | Cervical cancer | QIAscreen HPV PCR Test | Netherlands Cancer Institute |
| 0710501 | Melanoma | MSI analysis system | Erasmus MC |
| 0740103 | Colorectal Cancer | MSI analysis system | Netherlands Cancer Institute |
| 0750103 | Cervical cancer | CDKN2A/p16 IHC | Netherlands Cancer Institute |
| 0750103 | Cervical cancer | QIAscreen HPV PCR Test | Netherlands Cancer Institute |
| 0840201 | Kidney cancer | smMIP panel sequencing (RUO) | Radboudumc |
| 0870201 | Prostate cancer | Oncomine NGS gene-panel (custom) | Erasmus MC |
| 0920401 | Breast cancer | Oncomine NGS gene-panel (custom) | Erasmus MC |
| 1000412 | Prostate cancer | MSI analysis system | Erasmus MC |
| 1000431 | Melanoma | smMIP panel sequencing (RUO) | Radboudumc |
| 1000502 | Colorectal Cancer | MSI analysis system | Erasmus MC |
| 1000731 | Colorectal Cancer | MSI analysis system | Erasmus MC |
| 1100521 | Colorectal cancer | Oncomine NGS gene-panel (custom) | Erasmus MC |
| 1100631 | Breast cancer | Archer FusionPlex Solid | Erasmus MC |
| 1140501 | Melanoma | Oncomine NGS gene-panel (custom) | Erasmus MC |
| 1190103 | Head-neck cancer | QIAscreen HPV PCR Test | Netherlands Cancer Institute |
| 1200611 | Colorectal Cancer | MSI analysis system | Erasmus MC |
| 1210601 | Breast cancer | HER2/neu FISH | University Medical Center Utrecht |
| 1280201 | Melanoma | Oncomine NGS gene-panel (custom) | Erasmus MC |
| 1280201 | Melanoma | Oncomine TML assay | Erasmus MC |
| 1300611 | Breast cancer | HER2/neu FISH | University Medical Center Utrecht |
| 1301103 | Bladder cancer | CDKN2A/p16 IHC | Netherlands Cancer Institute |
| 1320601 | Colorectal cancer | Oncomine NGS gene-panel (custom) | Erasmus MC |
| 1360103 | Vulva cancer | QIAscreen HPV PCR Test | Netherlands Cancer Institute |
| 1380101 | Cancer of unknown primary | Archer FusionPlex Solid | Erasmus MC |
| 1380103 | Lung cancer | CDKN2A/p16 IHC | Netherlands Cancer Institute |
| 1400701 | Bladder cancer | MSI analysis system | Erasmus MC |
| 1410801 | Esophageal cancer | Archer FusionPlex Solid | Erasmus MC |
| 1460201 | Mesothelioma | Archer FusionPlex Solid | Erasmus MC |
| 1480103 | Colorectal Cancer | MMR IHC / MLH1 MS-MLPA | Netherlands Cancer Institute |
| 1530701 | Glioma | Oncomine NGS gene-panel (custom) | Erasmus MC |
| 1570103 | Head-neck cancer | QIAscreen HPV PCR Test | Netherlands Cancer Institute |
| 1570201 | Melanoma | Oncomine TML assay | Erasmus MC |
| 1810501 | Melanoma | HER2/neu FISH | University Medical Center Utrecht |
| 1920103 | GI-tract | QIAscreen HPV PCR Test | Netherlands Cancer Institute |
| 1990103 | Ovarian cancer | CDKN2A/p16 IHC | Netherlands Cancer Institute |
| 2000202 | Breast cancer | smMIP panel sequencing (RUO) | Radboudumc |
| 2000502 | Colorectal Cancer | MSI analysis system | Erasmus MC |
| 2010401 | Colorectal Cancer | MSI analysis system | Erasmus MC |
| 2010431 | Melanoma | Oncomine NGS gene-panel (custom) | Erasmus MC |
| 2080101 | Lung cancer | Archer FusionPlex Solid | Erasmus MC |
| 2080201 | Colorectal cancer | Oncomine TML assay | Erasmus MC |

| SampleID | Tumor type | Validation test | Institute |
|---|---|---|---|
| 2100321 | Colorectal cancer | smMIP panel sequencing (RUO) | Radboudumc |
| 2110103 | Vaginal cancer | CDKN2A/p16 IHC | Netherlands Cancer Institute |
| 2110103 | Vaginal cancer | QIAscreen HPV PCR Test | Netherlands Cancer Institute |
| 2140103 | Bladder cancer | CDKN2A/p16 IHC | Netherlands Cancer Institute |
| 2200102 | Cervical cancer | MSI analysis system | Erasmus MC |
| 2210701 | Colorectal Cancer | MSI analysis system | Erasmus MC |
| 2280103 | Sarcoma | CDKN2A/p16 IHC | Netherlands Cancer Institute |
| 2300211 | Lymphoma | MSI analysis system | Erasmus MC |
| 2310011 | Breast cancer | Archer FusionPlex Solid | Erasmus MC |
| 2350103 | Penile cancer | QIAscreen HPV PCR Test | Netherlands Cancer Institute |
| 2350301 | Glioma | Oncomine NGS gene-panel (custom) | Erasmus MC |
| 2420801 | Melanoma | Oncomine NGS gene-panel (custom) | Erasmus MC |
| 2510211 | Lung cancer | Archer FusionPlex Solid | Erasmus MC |
| 2510211 | Lung cancer | Oncomine NGS gene-panel (custom) | Erasmus MC |
| 2580103 | Lung cancer | CDKN2A/p16 IHC | Netherlands Cancer Institute |
| 2580201 | Liver cancer | Oncomine TML assay | Erasmus MC |
| 2600401 | Colorectal Cancer | MSI analysis system | Erasmus MC |
| 2720103 | Stomach cancer | QIAscreen HPV PCR Test | Netherlands Cancer Institute |
| 2780103 | Anal cancer | CDKN2A/p16 IHC | Netherlands Cancer Institute |
| 2800701 | Prostate cancer | smMIP panel sequencing (RUO) | Radboudumc |
| 2810103 | Vulva cancer | QIAscreen HPV PCR Test | Netherlands Cancer Institute |
| 2900103 | Neuroendocrine | QIAscreen HPV PCR Test | Netherlands Cancer Institute |
| 3000421 | Breast cancer | HER2/neu FISH | University Medical Center Utrecht |
| 3000502 | Glioma | MSI analysis system | Erasmus MC |
| 3010601 | Colorectal Cancer | MSI analysis system | Erasmus MC |
| 3101103 | Cervical cancer | QIAscreen HPV PCR Test | Netherlands Cancer Institute |
| 3110501 | Colorectal Cancer | MSI analysis system | Erasmus MC |
| 3120103 | Stomach cancer | QIAscreen HPV PCR Test | Netherlands Cancer Institute |
| 3130103 | Lung cancer | CDKN2A/p16 IHC | Netherlands Cancer Institute |
| 3150103 | Lung cancer | MET DISH | Netherlands Cancer Institute |
| 3190101 | Lung cancer | Archer FusionPlex Solid | Erasmus MC |
| 3190103 | Sarcoma | CDKN2A/p16 IHC | Netherlands Cancer Institute |
| 3200111 | Breast cancer | HER2/neu FISH | University Medical Center Utrecht |
| 3220601 | Colorectal cancer | Oncomine NGS gene-panel (custom) | Erasmus MC |
| 3250101 | Breast cancer | smMIP panel sequencing (RUO) | Radboudumc |
| 3300211 | Breast cancer | HER2/neu FISH | University Medical Center Utrecht |
| 3330103 | Lung cancer | CDKN2A/p16 IHC | Netherlands Cancer Institute |
| 3350101 | Colorectal cancer | MSI analysis system | Erasmus MC |
| 3350101 | Colorectal cancer | smMIP panel sequencing (RUO) | Radboudumc |
| 3430201 | Lung cancer | Archer FusionPlex Solid | Erasmus MC |
| 3501103 | Colorectal cancer | MET DISH | Netherlands Cancer Institute |
| 3660101 | Bladder cancer | HER2/neu FISH | University Medical Center Utrecht |
| 3700111 | Colorectal cancer | Oncomine NGS gene-panel (custom) | Erasmus MC |
| 3740201 | Head and Neck cancer | smMIP panel sequencing (RUO) | Radboudumc |
| 3790103 | Vulva cancer | QIAscreen HPV PCR Test | Netherlands Cancer Institute |
| 3800401 | Pancreatic cancer | smMIP panel sequencing (RUO) | Radboudumc |
| 3800431 | Melanoma | Oncomine NGS gene-panel (custom) | Erasmus MC |
| 3900501 | Colorectal Cancer | MSI analysis system | Erasmus MC |
| 3901103 | Lung cancer | MET DISH | Netherlands Cancer Institute |
| 3990101 | Penile cancer | QIAscreen HPV PCR Test | Netherlands Cancer Institute |
| 4000901 | Colorectal Cancer | MSI analysis system | Erasmus MC |
| 4010501 | Colorectal Cancer | MSI analysis system | Erasmus MC |
| 4060103 | Cancer of unknown primary | CDKN2A/p16 IHC | Netherlands Cancer Institute |
| 4100102 | Cancer of unknown primary | MSI analysis system | Erasmus MC |
| 4100511 | Breast cancer | Archer FusionPlex Solid | Erasmus MC |
| 4100811 | Cholangiocarcinoma | smMIP panel sequencing (RUO) | Radboudumc |
| 4160103 | Cancer of unknown primary | CDKN2A/p16 IHC | Netherlands Cancer Institute |
| 4180103 | Lung cancer | MET DISH | Netherlands Cancer Institute |
| 4201103 | Lung cancer | CDKN2A/p16 IHC | Netherlands Cancer Institute |
| 4250101 | Lung cancer | smMIP panel sequencing (RUO) | Radboudumc |

| SampleID | Tumor type | Validation test | Institute |
|---|---|---|---|
| 4280103 | Penile cancer | CDKN2A/p16 IHC | Netherlands Cancer Institute |
| 4320801 | Skin cancer | Oncomine NGS gene-panel (custom) | Erasmus MC |
| 4330103 | Lung cancer | CDKN2A/p16 IHC | Netherlands Cancer Institute |
| 4410311 | Colorectal cancer | Oncomine NGS gene-panel (custom) | Erasmus MC |
| 4410501 | Gallbladder cancer | MSI analysis system | Erasmus MC |
| 4480201 | Melanoma | Oncomine TML assay | Erasmus MC |
| 4500401 | Lung cancer | Archer FusionPlex Solid | Erasmus MC |
| 4600103 | Esophageal cancer | QIAscreen HPV PCR Test | Netherlands Cancer Institute |
| 4610701 | Prostate cancer | MSI analysis system | Erasmus MC |
| 4640103 | Anus cancer | QIAscreen HPV PCR Test | Netherlands Cancer Institute |
| 4730103 | Sarcoma | CDKN2A/p16 IHC | Netherlands Cancer Institute |
| 4800801 | Colorectal Cancer | MSI analysis system | Erasmus MC |
| 4900321 | Breast cancer | Archer FusionPlex Solid | Erasmus MC |
| 4920401 | Prostate cancer | Oncomine NGS gene-panel (custom) | Erasmus MC |
| 4980101 | Lung cancer | Archer FusionPlex Solid | Erasmus MC |
| 4980101 | Lung cancer | Oncomine NGS gene-panel (custom) | Erasmus MC |
| 4980102 | Lung cancer | Archer FusionPlex Solid | Erasmus MC |
| 5000702 | Gastrointestinal stromal tumor | smMIP panel sequencing (RUO) | Radboudumc |
| 5100702 | Prostate cancer | MSI analysis system | Erasmus MC |
| 5110501 | Colorectal Cancer | MSI analysis system | Erasmus MC |
| 5120401 | Lung cancer | Archer FusionPlex Solid | Erasmus MC |
| 5120401 | Lung cancer | Archer FusionPlex Solid | Erasmus MC |
| 5180201 | Cancer of unknown primary | Oncomine TML assay | Erasmus MC |
| 5200102 | Colorectal Cancer | MSI analysis system | Erasmus MC |
| 5200221 | Colorectal cancer | HER2/neu FISH | University Medical Center Utrecht |
| 5200331 | Colorectal Cancer | MSI analysis system | Erasmus MC |
| 5220103 | Testis cancer | QIAscreen HPV PCR Test | Netherlands Cancer Institute |
| 5230301 | Sarcoma | smMIP panel sequencing (RUO) | Radboudumc |
| 5311103 | Vaginal cancer | QIAscreen HPV PCR Test | Netherlands Cancer Institute |
| 5400121 | Esophageal cancer | Oncomine NGS gene-panel (custom) | Erasmus MC |
| 5490103 | Endometrial Cancer | MMR IHC / MLH1 MS-MLPA | Netherlands Cancer Institute |
| 5530103 | Esophageal cancer | QIAscreen HPV PCR Test | Netherlands Cancer Institute |
| 5550101 | Breast cancer | HER2/neu FISH | University Medical Center Utrecht |
| 5580103 | Lung cancer | MET DISH | Netherlands Cancer Institute |
| 5600411 | Prostate cancer | Oncomine NGS gene-panel (custom) | Erasmus MC |
| 5640201 | Gastrointestinal stromal tumor | smMIP panel sequencing (RUO) | Radboudumc |
| 5700103 | Lung Cancer | MMR IHC / MLH1 MS-MLPA | Netherlands Cancer Institute |
| 5700111 | Neuroendocrine | Oncomine NGS gene-panel (custom) | Erasmus MC |
| 5710103 | Lung cancer | CDKN2A/p16 IHC | Netherlands Cancer Institute |
| 5720103 | Anus cancer | QIAscreen HPV PCR Test | Netherlands Cancer Institute |
| 5730501 | Colorectal cancer | Oncomine NGS gene-panel (custom) | Erasmus MC |
| 5740201 | Breast cancer | smMIP panel sequencing (RUO) | Radboudumc |
| 5760103 | Lung cancer | MET DISH | Netherlands Cancer Institute |
| 5801103 | Lung cancer | MET DISH | Netherlands Cancer Institute |
| 5870201 | Melanoma | Oncomine NGS gene-panel (custom) | Erasmus MC |
| 5940101 | Melanoma | smMIP panel sequencing (RUO) | Radboudumc |
| 5950103 | Anal cancer | CDKN2A/p16 IHC | Netherlands Cancer Institute |
| 6000321 | Colorectal cancer | MSI analysis system | Erasmus MC |
| 6000321 | Colorectal cancer | smMIP panel sequencing (RUO) | Radboudumc |
| 6030103 | Lung cancer | CDKN2A/p16 IHC | Netherlands Cancer Institute |
| 6060103 | Colorectal Cancer | MMR IHC / MLH1 MS-MLPA | Netherlands Cancer Institute |
| 6100121 | Pancreatic cancer | MSI analysis system | Erasmus MC |
| 6100131 | Stomach cancer | Oncomine NGS gene-panel (custom) | Erasmus MC |
| 6100331 | Lymphoma | smMIP panel sequencing (RUO) | Radboudumc |
| 6110501 | Head and Neck cancer | smMIP panel sequencing (RUO) | Radboudumc |
| 6120103 | Ovarian cancer | CDKN2A/p16 IHC | Netherlands Cancer Institute |
| 6130103 | Lung cancer | CDKN2A/p16 IHC | Netherlands Cancer Institute |
| 6200102 | Colorectal Cancer | MSI analysis system | Erasmus MC |
| 6200103 | Lung cancer | MET DISH | Netherlands Cancer Institute |
| 6200511 | Lymphoma | Oncomine NGS gene-panel (custom) | Erasmus MC |

| SampleID | Tumor type | Validation test | Institute |
|---|---|---|---|
| 6220401 | Colorectal cancer | Oncomine NGS gene-panel (custom) | Erasmus MC |
| 6250101 | Melanoma | smMIP panel sequencing (RUO) | Radboudumc |
| 6250201 | Esophageal cancer | MSI analysis system | Erasmus MC |
| 6260103 | Lung cancer | MET DISH | Netherlands Cancer Institute |
| 6280201 | Melanoma | Oncomine TML assay | Erasmus MC |
| 6360103 | Cancer of unknown primary | CDKN2A/p16 IHC | Netherlands Cancer Institute |
| 6360103 | Cancer of unknown primary | MET DISH | Netherlands Cancer Institute |
| 6370201 | Melanoma | Oncomine NGS gene-panel (custom) | Erasmus MC |
| 6420401 | Melanoma | Oncomine NGS gene-panel (custom) | Erasmus MC |
| 6520601 | Melanoma | Oncomine NGS gene-panel (custom) | Erasmus MC |
| 6530103 | Sarcoma | CDKN2A/p16 IHC | Netherlands Cancer Institute |
| 6560103 | Lung cancer | CDKN2A/p16 IHC | Netherlands Cancer Institute |
| 6600103 | GI-tract | QIAscreen HPV PCR Test | Netherlands Cancer Institute |
| 6601103 | Cervical cancer | QIAscreen HPV PCR Test | Netherlands Cancer Institute |
| 6690101 | Lung cancer | Archer FusionPlex Solid | Erasmus MC |
| 6770101 | Lung cancer | Oncomine NGS gene-panel (custom) | Erasmus MC |
| 6770102 | Lung cancer | Oncomine NGS gene-panel (custom) | Erasmus MC |
| 6840201 | Colorectal Cancer | MSI analysis system | Erasmus MC |
| 6900601 | Colorectal Cancer | MSI analysis system | Erasmus MC |
| 6901103 | Cancer of unknown primary | CDKN2A/p16 IHC | Netherlands Cancer Institute |
| 6920103 | Cervical cancer | QIAscreen HPV PCR Test | Netherlands Cancer Institute |
| 6930103 | Liver cancer | QIAscreen HPV PCR Test | Netherlands Cancer Institute |
| 6930501 | Glioma | Oncomine NGS gene-panel (custom) | Erasmus MC |
| 6960101 | Skin cancer | QIAscreen HPV PCR Test | Netherlands Cancer Institute |
| 7000103 | Cervical cancer | CDKN2A/p16 IHC | Netherlands Cancer Institute |
| 7000103 | Cervical cancer | QIAscreen HPV PCR Test | Netherlands Cancer Institute |
| 7000321 | Unknown | smMIP panel sequencing (RUO) | Radboudumc |
| 7000711 | Stomach cancer | HER2/neu FISH | University Medical Center Utrecht |
| 7021103 | Colorectal Cancer | MSI analysis system | Netherlands Cancer Institute |
| 7050201 | Colorectal Cancer | MSI analysis system | Erasmus MC |
| 7060103 | Lung cancer | MET DISH | Netherlands Cancer Institute |
| 7100031 | Colorectal Cancer | MSI analysis system | Erasmus MC |
| 7100103 | Head-neck cancer | QIAscreen HPV PCR Test | Netherlands Cancer Institute |
| 7100131 | Lymphoma | Oncomine NGS gene-panel (custom) | Erasmus MC |
| 7100331 | Breast cancer | HER2/neu FISH | University Medical Center Utrecht |
| 7190101 | Lung cancer | Archer FusionPlex Solid | Erasmus MC |
| 7201103 | Head and Neck cancer | CDKN2A/p16 IHC | Netherlands Cancer Institute |
| 7201103 | Head-neck cancer | QIAscreen HPV PCR Test | Netherlands Cancer Institute |
| 7280101 | Colorectal cancer | Oncomine NGS gene-panel (custom) | Erasmus MC |
| 7280201 | Prostate cancer | Oncomine TML assay | Erasmus MC |
| 7310211 | Breast cancer | Oncomine NGS gene-panel (custom) | Erasmus MC |
| 7330103 | Cancer of unknown primary | CDKN2A/p16 IHC | Netherlands Cancer Institute |
| 7330501 | Breast cancer | Archer FusionPlex Solid | Erasmus MC |
| 7380103 | Lung cancer | MET DISH | Netherlands Cancer Institute |
| 7410103 | Head-neck cancer | QIAscreen HPV PCR Test | Netherlands Cancer Institute |
| 7411103 | Head-neck cancer | QIAscreen HPV PCR Test | Netherlands Cancer Institute |
| 7480101 | Lung cancer | Oncomine NGS gene-panel (custom) | Erasmus MC |
| 7600601 | Kidney cancer | smMIP panel sequencing (RUO) | Radboudumc |
| 7650101 | Lung cancer | Archer FusionPlex Solid | Erasmus MC |
| 7700801 | Colorectal Cancer | MSI analysis system | Erasmus MC |
| 7780103 | Colorectal Cancer | MMR IHC / MLH1 MS-MLPA | Netherlands Cancer Institute |
| 7870201 | Kidney cancer | Oncomine NGS gene-panel (custom) | Erasmus MC |
| 7930501 | Colorectal cancer | Oncomine NGS gene-panel (custom) | Erasmus MC |
| 7990103 | Cancer of unknown primary | CDKN2A/p16 IHC | Netherlands Cancer Institute |
| 8000031 | Breast cancer | smMIP panel sequencing (RUO) | Radboudumc |
| 8000702 | Unknown | MSI analysis system | Erasmus MC |
| 8000811 | Colorectal Cancer | MSI analysis system | Erasmus MC |
| 8100102 | Colorectal Cancer | MSI analysis system | Erasmus MC |
| 8100111 | Esophageal cancer | smMIP panel sequencing (RUO) | Radboudumc |
| 8110501 | Colorectal Cancer | MSI analysis system | Erasmus MC |

| SampleID | Tumor type | Validation test | Institute |
|---|---|---|---|
| 8130301 | Stomach cancer | MSI analysis system | Erasmus MC |
| 8210501 | Colorectal Cancer | MSI analysis system | Erasmus MC |
| 8230301 | Colorectal Cancer | MSI analysis system | Erasmus MC |
| 8250301 | Colorectal cancer | Oncomine NGS gene-panel (custom) | Erasmus MC |
| 8340101 | Neuroendocrine tumor | smMIP panel sequencing (RUO) | Radboudumc |
| 8380103 | Lung cancer | CDKN2A/p16 IHC | Netherlands Cancer Institute |
| 8460103 | Lung cancer | MET DISH | Netherlands Cancer Institute |
| 8600431 | Endometrial cancer | Oncomine NGS gene-panel (custom) | Erasmus MC |
| 8630103 | Lung cancer | MET DISH | Netherlands Cancer Institute |
| 8700401 | Breast cancer | HER2/neu FISH | University Medical Center Utrecht |
| 8720501 | Colorectal cancer | HER2/neu FISH | University Medical Center Utrecht |
| 8730103 | Lung cancer | MET DISH | Netherlands Cancer Institute |
| 8740201 | Breast cancer | HER2/neu FISH | University Medical Center Utrecht |
| 8740201 | Breast cancer | smMIP panel sequencing (RUO) | Radboudumc |
| 8750103 | Cancer of unknown primary | MMR IHC / MLH1 MS-MLPA | Netherlands Cancer Institute |
| 8970103 | Lung cancer | MET DISH | Netherlands Cancer Institute |
| 8980101 | Colorectal cancer | QIAscreen HPV PCR Test | Netherlands Cancer Institute |
| 8980103 | Colorectal Cancer | MMR IHC / MLH1 MS-MLPA | Netherlands Cancer Institute |
| 9001103 | Lung cancer | MET DISH | Netherlands Cancer Institute |
| 9010211 | Melanoma | Archer FusionPlex Solid | Erasmus MC |
| 9010211 | Melanoma | Oncomine NGS gene-panel (custom) | Erasmus MC |
| 9100811 | Unknown | smMIP panel sequencing (RUO) | Radboudumc |
| 9110103 | Cancer of unknown primary | CDKN2A/p16 IHC | Netherlands Cancer Institute |
| 9110211 | Colorectal cancer | Oncomine NGS gene-panel (custom) | Erasmus MC |
| 9180201 | Melanoma | Oncomine NGS gene-panel (custom) | Erasmus MC |
| 9180201 | Melanoma | Oncomine TML assay | Erasmus MC |
| 9200111 | Breast cancer | HER2/neu FISH | University Medical Center Utrecht |
| 9200721 | Lung cancer | Oncomine NGS gene-panel (custom) | Erasmus MC |
| 9320501 | Pancreatic cancer | Archer FusionPlex Solid | Erasmus MC |
| 9330103 | Breast Cancer | CDKN2A/p16 IHC | Netherlands Cancer Institute |
| 9360103 | Cervical cancer | QIAscreen HPV PCR Test | Netherlands Cancer Institute |
| 9501103 | Cervical cancer | QIAscreen HPV PCR Test | Netherlands Cancer Institute |
| 9520103 | Lung cancer | MET DISH | Netherlands Cancer Institute |
| 9600103 | Breast Cancer | MMR IHC / MLH1 MS-MLPA | Netherlands Cancer Institute |
| 9620401 | Colorectal cancer | Oncomine NGS gene-panel (custom) | Erasmus MC |
| 9650101 | Esophageal cancer | MSI analysis system | Erasmus MC |
| 9700801 | Melanoma | smMIP panel sequencing (RUO) | Radboudumc |
| 9710501 | Stomach cancer | MSI analysis system | Erasmus MC |
| 9750101 | Colorectal Cancer | MSI analysis system | Erasmus MC |
| 9950103 | Anal cancer | CDKN2A/p16 IHC | Netherlands Cancer Institute |
| 9950103 | Anus cancer | QIAscreen HPV PCR Test | Netherlands Cancer Institute |
