## Supplementary Table 2 for "Clinical validation of Whole Genome Sequencing for cancer diagnostics"

**WGS sensitivity model considering:**

| | |
|---|---|
| Tumor ploidy | 2 |
| Variant copy number | 1 |
| x (Required reads) | 5 |
| purity\coverage | 100 |

| **Tumor purity** | **Sensitivity** |
|---|---|
| **0.19** | **95.97** |

Model details per 0.05 purity and coverage

| purity\coverage | 5 | 10 | 15 | 20 | 25 | 30 | 35 | 40 | 50 | 60 | 70 | 80 | 90 | 100 | 120 | 150 | 200 | 250 | 300 |
|---|---|---|---|---|---|---|---|---|---|---|---|---|---|---|---|---|---|---|---|
| 0.05 | 0.00 | 0.00 | 0.00 | 0.02 | 0.05 | 0.11 | 0.21 | 0.37 | 0.91 | 1.86 | 3.29 | 5.27 | 7.80 | 10.88 | 18.47 | 32.25 | 55.95 | 74.70 | 86.79 |
| 0.1 | 0.00 | 0.02 | 0.11 | 0.37 | 0.91 | 1.86 | 3.29 | 5.27 | 10.88 | 18.47 | 27.46 | 37.12 | 46.79 | 55.95 | 71.49 | 86.79 | 97.07 | 99.47 | 99.91 |
| 0.15 | 0.00 | 0.11 | 0.60 | 1.86 | 4.21 | 7.80 | 12.61 | 18.47 | 32.25 | 46.79 | 60.22 | 71.49 | 80.30 | 86.79 | 94.50 | 98.72 | 99.91 | 100.00 | 100.00 |
| 0.2 | 0.02 | 0.37 | 1.86 | 5.27 | 10.88 | 18.47 | 27.46 | 37.12 | 55.95 | 71.49 | 82.70 | 90.04 | 94.50 | 97.07 | 99.24 | 99.91 | 100.00 | 100.00 | 100.00 |
| 0.25 | 0.05 | 0.91 | 4.21 | 10.88 | 20.62 | 32.25 | 44.40 | 55.95 | 74.70 | 86.79 | 93.60 | 97.07 | 98.72 | 99.47 | 99.91 | 100.00 | 100.00 | 100.00 | 100.00 |
| 0.3 | 0.11 | 1.86 | 7.80 | 18.47 | 32.25 | 46.79 | 60.22 | 71.49 | 86.79 | 94.50 | 97.89 | 99.24 | 99.74 | 99.91 | 99.99 | 100.00 | 100.00 | 100.00 | 100.00 |
| 0.35 | 0.21 | 3.29 | 12.61 | 27.46 | 44.40 | 60.22 | 73.13 | 82.70 | 93.60 | 97.89 | 99.36 | 99.82 | 99.95 | 99.99 | 100.00 | 100.00 | 100.00 | 100.00 | 100.00 |
| 0.4 | 0.37 | 5.27 | 18.47 | 37.12 | 55.95 | 71.49 | 82.70 | 90.04 | 97.07 | 99.24 | 99.82 | 99.96 | 99.99 | 100.00 | 100.00 | 100.00 | 100.00 | 100.00 | 100.00 |
| 0.45 | 0.60 | 7.80 | 25.12 | 46.79 | 66.16 | 80.30 | 89.30 | 94.50 | 98.72 | 99.74 | 99.95 | 99.99 | 100.00 | 100.00 | 100.00 | 100.00 | 100.00 | 100.00 | 100.00 |
| 0.5 | 0.91 | 10.88 | 32.25 | 55.95 | 74.70 | 86.79 | 93.60 | 97.07 | 99.47 | 99.91 | 99.99 | 100.00 | 100.00 | 100.00 | 100.00 | 100.00 | 100.00 | 100.00 | 100.00 |
| 0.55 | 1.33 | 14.46 | 39.56 | 64.25 | 81.53 | 91.38 | 96.28 | 98.49 | 99.78 | 99.97 | 100.00 | 100.00 | 100.00 | 100.00 | 100.00 | 100.00 | 100.00 | 100.00 | 100.00 |
| 0.6 | 1.86 | 18.47 | 46.79 | 71.49 | 86.79 | 94.50 | 97.89 | 99.24 | 99.91 | 99.99 | 100.00 | 100.00 | 100.00 | 100.00 | 100.00 | 100.00 | 100.00 | 100.00 | 100.00 |
| 0.65 | 2.51 | 22.83 | 53.73 | 77.63 | 90.73 | 96.56 | 98.83 | 99.63 | 99.97 | 100.00 | 100.00 | 100.00 | 100.00 | 100.00 | 100.00 | 100.00 | 100.00 | 100.00 | 100.00 |
| 0.7 | 3.29 | 27.46 | 60.22 | 82.70 | 93.60 | 97.89 | 99.36 | 99.82 | 99.99 | 100.00 | 100.00 | 100.00 | 100.00 | 100.00 | 100.00 | 100.00 | 100.00 | 100.00 | 100.00 |
| 0.75 | 4.21 | 32.25 | 66.16 | 86.79 | 95.64 | 98.72 | 99.66 | 99.91 | 100.00 | 100.00 | 100.00 | 100.00 | 100.00 | 100.00 | 100.00 | 100.00 | 100.00 | 100.00 | 100.00 |
| 0.8 | 5.27 | 37.12 | 71.49 | 90.04 | 97.07 | 99.24 | 99.82 | 99.96 | 100.00 | 100.00 | 100.00 | 100.00 | 100.00 | 100.00 | 100.00 | 100.00 | 100.00 | 100.00 | 100.00 |
| 0.85 | 6.46 | 41.99 | 76.20 | 92.56 | 98.06 | 99.55 | 99.91 | 99.98 | 100.00 | 100.00 | 100.00 | 100.00 | 100.00 | 100.00 | 100.00 | 100.00 | 100.00 | 100.00 | 100.00 |
| 0.9 | 7.80 | 46.79 | 80.30 | 94.50 | 98.72 | 99.74 | 99.95 | 99.99 | 100.00 | 100.00 | 100.00 | 100.00 | 100.00 | 100.00 | 100.00 | 100.00 | 100.00 | 100.00 | 100.00 |
| 0.95 | 9.28 | 51.46 | 83.81 | 95.97 | 99.17 | 99.85 | 99.98 | 100.00 | 100.00 | 100.00 | 100.00 | 100.00 | 100.00 | 100.00 | 100.00 | 100.00 | 100.00 | 100.00 | 100.00 |
| 1 | 10.88 | 55.95 | 86.79 | 97.07 | 99.47 | 99.91 | 99.99 | 100.00 | 100.00 | 100.00 | 100.00 | 100.00 | 100.00 | 100.00 | 100.00 | 100.00 | 100.00 | 100.00 | 100.00 |

---

**Goal**

The goal is to use a simple model to estimate what percentage of the non-subclonal mutations of a sample of tumor cells we should detect for various combinations of purity, ploidy, coverage, distribution of variant copy numbers, and number of reads required to be called as a variant.

**Definitions**

Consider a sample that is a mixture of germline and tumor cells.
Define the purity P as the (exact) percentage of the cells that is a tumor cell.
Suppose the following:
-The sample has been sequenced.
-All tumor cells are genetically identical.
-All germline cells are genetically identical.
-The germline genome is known precisely.
-All mutations in the tumor genome wrt the germline genome are SNV's.
-The tumor ploidy is equal to M.
-All reads of the tumor sample have the exact same length L.

Define C as the average coverage of the tumor sample.
Let N be the number of variants present in the tumor genome wrt the germline genome, and label these variants v_1, v_2,..., v_N.
Let V_i be the variant vopy number of the variant v_i, and let r_i be the number of reads required to call v_i as a variant, for i=1,...,N.

**Model**

Let X be the number of variants that we actually detect. Now what is the probability distribution of X? What is the expectation of X?

Define Y_i as "v_i was detected" and Z_i as "the number of reads of v_i found from the tumor sample sequencing" for i=1,...,N.
This means that Y_i is equivalent to "Z_i>=r_i", so Y_i is a Bernoulli stochastic variable with succes probability q_i=P(Z_i>=r_i).

Since X is by definition the number of Y_i's that are a succes, it might be tempting to say that X is binomially distributed.
There is a potential problem that the Y_i's are not independent, strictly speaking, because all of the reads are taken from the same pool of DNA from the tumor sample.
Fortunately, this interdependence is weak enough that assuming independence can still be reasonably justified.
Since the q_i are not necessarily all identical, X is actually Poisson binomially distributed with probabilities q_1,...,q_N.
This implies that E(X)=sum(q_i) and Var(X)=sum(q_i(1-q_i))

The proportion of the existing variants that we actually detect is equal to X/N, and we see that
E(X/N)=sum(q_i)/N and Var(X/N)=sum(q_i(1-q_i))/N^2

Note that independence of the Y_i is not required to determine the formula for the expectation, since X=sum(1_{Y_i}) would still be true without this assumption.

For this model to be useful, we need to be able to calculate or approximate the probabilities q_i=P(Z_i>=r_i).
The derivation below explains why Poisson(P*V_i*C/(P*(M-2)+2)) is a reasonable approximation to the distribution of Z_i.

**Poisson Derivation**

Each read from the sequencing is independently chosen out of all of the pieces of DNA of appropriate length in the prepared sample.
A haploid human genome contains c=3.1 billion base pairs. This means that in the sequencing of the tumor genome, the total number of reads was C*c/L, so at least a few million.
Then Z_i is binomially distributed, let's say bin(n_i,p_i). Then n_i=C*c/L and p_i="percentage of total reads that include variant v_i"
In the genome there are L spots where a read could start to include the position of v_i. If we let N_c be the number of cells in the tumor sample, then
p_i="number of positions in all of the DNA in the tumor sample where a read can start, if it needs to contains v_i" / "number of positions in all of the DNA in the tumor sample where a read can start"
="number of tumor cells" * "number of positions in the DNA of a tumor cell where reads containing v_i can start" / "total number of bases in all of the DNA of all of the cells in the tumor sample, combined"
=(N_c*P*V_i*L)/(N_c*P*M*c+N_c*(1-P)*2*c)=P*V_i*L/(c*(P*(M-2)+2))
p_i is a very small number.
Since n_i is so large and p_i is so small, the distribution of Z_i can reasonably be approximated by a Poission(l_i) distribution with l_i=n_i*p_i=P*V_i*C/(P*(M-2)+2).
