## Supplementary Table 3 for "Clinical validation of Whole Genome Sequencing for cancer diagnostics"

| Sample | WGS somatic aberrations | | | Run 1 | Run 2 |
|---|---|---|---|---|---|
| **Tumor A** | **Gene** | **Coding** | **Protein** | **Variant Freq** | **Variant Freq** |
| | *CTNNB1* | c.94G>C | p.Asp32His | 77% | 83% |
| | *ESR1* | c.667G>A | p.Ala223Thr | 19% | 17% |
| | *TP53* | c.746G>T | p.Arg249Met | 83% | 79% |
| | *FANCG* | | copy-gain | 7 | 7 |
| | *RB1* | | copy-loss | 0 | 0 |
| | Microsatellite Status | | stable | 0.241 | 0.245 |
| | Mutational Load | | high | 213 | 219 |
| **Sample** | **WGS somatic aberrations** | | | **Run 1** | **Run 2** |
| **Tumor B** | **Gene** | **Coding** | **Protein** | **Variant Freq** | **Variant Freq** |
| | *ERBB4* | c.3493C>T | p.Pro1165Ser | 11% | 15% |
| | *PIK3CB* | c.77C>T | p.Ser26Phe | 26% | 20% |
| | *KDR* | c.3482G>A | p.Gly1161Glu | 22% | 15% |
| | *APC* | c.7427G>A | p.Arg2476Lys | 16% | 21% |
| | *ESR1* | c.652G>A | p.Asp218Asn | 27% | 17% |
| | *PMS2* | c.1439G>A | p.Gly480Glu | 22% | 23% |
| | *CDKN2A* | c.151-1G>A | | 28% | 24% |
| | *NOTCH1* | c.3042_3043delGGinsAA | p.Gly1015Ser | 20% | 28% |
| | *NOTCH1* | c.1607C>T | p.Pro536Leu | 13% | nd (10%)* |
| | *CTNNA3* | c.1666G>A | p.Asp556Asn | 17% | 14% |
| | *FLT3* | c.1172C>T | p.Pro391Leu | 7% | 13% |
| | *TSC2* | c.4591G>C | p.Val1531Leu | 21% | 22% |
| | *TP53* | c.404G>T | p.Cys135Phe | 26% | 23% |
| | *TP53* | c.96+1G>T | | 18% | 16% |
| | *SMARCB1* | c.740C>T | p.Ser247Phe | 18% | 19% |
| | Microsatellite Status | | stable | 0.11 | 0.11 |
| | Mutational Load | | high | 1167 | 1171 |

**NOTCH* p.Pro536Leu was detected in run 2 with 10% VAF but filtered out due to stringent quality criteria.
