## Supplementary Table 4 for "Clinical validation of Whole Genome Sequencing for cancer diagnostics"

| nr | Tumor | pathological TCP (pTCP) pre-DNAiso section | post-DNAiso section | Average | Difference | molecular purity (mTCP) | Difference pTCP (average) vs. mTCP |
|---|---|---|---|---|---|---|---|
| 1 | colorectal | 35 | 35 | 35 | 0 | 60 | 25 |
| 2 | pancreas | 70 | 50 | 60 | -20 | 27 | -33 |
| 3 | ovary | 70 | 40 | 55 | -30 | 25 | -30 |
| 4 | endomerium | 80 | 50 | 65 | -30 | 98 | 33 |
| 5 | lung | 50 | 10 | 30 | -40 | 25 | -5 |
| 6 | cervix | 70 | 70 | 70 | 0 | 30 | -40 |
| 7 | breast | 35 | 30 | 32.5 | -5 | 64 | 31.5 |
| 8 | breast | 70 | 30 | 50 | -40 | 18 | -32 |
| 9 | GI tract | 70 | 45 | 57.5 | -25 | 25 | -32.5 |
| 10 | ovary | 65 | 60 | 62.5 | -5 | 24 | -38.5 |
| 11 | renal cell carcinoma | 60 | 80 | 70 | 20 | 10 | -60 |
| 12 | colorectal | 70 | 70 | 70 | 0 | 35 | -35 |
| 13 | breast | 50 | 50 | 50 | 0 | 29 | -21 |
| 14 | lung | 30 | 75 | 52.5 | 45 | 18 | -34.5 |
| 15 | cervix | 80 | 80 | 80 | 0 | 18 | -62 |
| 16 | colorectal | 30 | 40 | 35 | 10 | 63 | 28 |
| 17 | melanoma | 80 | 80 | 80 | 0 | 41 | -39 |
| 18 | breast | 40 | 40 | 40 | 0 | 18 | -22 |
| 19 | melanoma | 80 | 60 | 70 | -20 | 36 | -34 |
| 20 | hepatocellular | 40 | 40 | 40 | 0 | 29 | -11 |
