## Supplementary Table 5 for "Clinical validation of Whole Genome Sequencing for cancer diagnostics"

| ID | Gene | coding | protein | type | WGS (tumor adjusted VAF) | Oncomine NGS panel VAF (coverage) |
|---|---|---|---|---|---|---|
| 0500121 | APC | c.4661delA | p.Thr1556fs | INDEL | 100% | covered, not called |
| 0700111 | APC | c.4012C>T | p.Gln1338* | SNP | 77% | 17% (791) |
| 0700111 | APC | c.2679_2686delAGTGTCAG | p.Glu893fs | INDEL | 44% | covered, not called |
| 1320601 | APC | c.4469_4470insG | p.His1490fs | INDEL | 78% | 17% (864) |
| 3220601 | APC | c.4661delA | p.Thr1556fs | INDEL | 33% | covered, not called |
| 3700111 | APC | c.4660_4661insA | p.Thr1556fs | INDEL | 43% | covered, not called |
| 4320801 | APC | c.3340C>T | p.Arg1114* | SNP | 68% | 50% (948) |
| 5700111 | APC | c.3766C>T | p.Gln1256* | SNP | 40% | 40% (1351) |
| 5700111 | APC | c.3921_3925delAAAAG | p.Glu1309fs | INDEL | 50% | 30% (735) |
| 7930501 | APC | c.4135G>T | p.Glu1379* | SNP | 30% | 27% (834) |
| 9620401 | APC | c.4348C>T | p.Arg1450* | SNP | 56% | 30% (283) |
| 9620401 | APC | c.2626C>T | p.Arg876* | SNP | 46% | 23% (811) |
| 0920401 | BRAF | c.1780G>A | p.Asp594Asn | SNP | 37% | 24% (1550) |
| 1100521 | BRAF | c.1799T>A | p.Val600Glu | SNP | 34% | 14% (923) |
| 1140501 | BRAF | c.1799T>A | p.Val600Glu | SNP | 68% | 50% (3398) |
| 1280201 | BRAF | c.1799T>A | p.Val600Glu | SNP | 59% | 38% (1425) |
| 2010431 | BRAF | c.1798_1799delGTinsAA | p.Val600Lys | INDEL | 76% | 8% (771) |
| 2350301 | BRAF | c.2285C>T | p.Ala762Val | SNP | 57% | not covered |
| 4410311 | BRAF | c.1799T>A | p.Val600Glu | SNP | 60% | 46% (2460) |
| 4920401 | BRAF | c.1801A>G | p.Lys601Glu | SNP | 67% | 43% (2140) |
| 4980101 | BRAF | c.1799T>A | p.Val600Glu | SNP | 15% | 16% (1234) |
| 5600411 | BRAF | c.1801A>G | p.Lys601Glu | SNP | 31% | 22% (1387) |
| 5730501 | BRAF | c.1780G>A | p.Asp594Asn | SNP | 48% | 43% (3747) |
| 5870201 | BRAF | c.1782T>G | p.Asp594Glu | SNP | 55% | 46% (1819) |
| 6200511 | BRAF | c.1780G>A | p.Asp594Asn | SNP | 65% | 30% (908) |
| 6220401 | BRAF | c.1799T>A | p.Val600Glu | SNP | 30% | 28% (1908) |
| 6370201 | BRAF | c.1798_1799delGTinsAA | p.Val600Lys | INDEL | 34% | 8% (463) |
| 6420401 | BRAF | c.1799T>A | p.Val600Glu | SNP | 40% | 16% (647) |
| 6520601 | BRAF | c.1799T>A | p.Val600Glu | SNP | 71% | 62% (3032) |
| 7100131 | BRAF | c.1803A>T | p.Lys601Asn | SNP | 41% | 40% (1266) |
| 7280101 | BRAF | c.1208delC | p.Pro403fs | INDEL | 96% | not covered |
| 7280101 | BRAF | c.1799T>A | p.Val600Glu | SNP | 100% | 7% (310) |
| 7310211 | BRAF | c.1406G>C | p.Gly469Ala | SNP | not called | 2% (60) |
| 7480101 | BRAF | c.1406G>T | p.Gly469Val | SNP | 44% | 37% (1791) |
| 7870201 | BRAF | c.1799_1801delTGA | p.Val600_Lys601delinsGlu | INDEL | 42% | 43% (1852) |
| 8250301 | BRAF | c.1799T>A | p.Val600Glu | SNP | 24% | 22% (1251) |
| 9010211 | BRAF | c.1406G>C | p.Gly469Ala | SNP | 73% | 35% (983) |
| 9110211 | BRAF | c.1799T>A | p.Val600Glu | SNP | 47% | 34% (2091) |
| 9180201 | BRAF | c.1798_1799delGTinsAA | p.Val600Lys | INDEL | 76% | 14% (1349) |
| 9200721 | BRAF | c.1780G>C | p.Asp594His | SNP | 50% | 38% (38) |
| 0500831 | CDKN2A | c.250G>T | p.Asp84Tyr | SNP | 100% | 70% (2688) |
| 1140501 | CDKN2A | c.341C>T | p.Pro114Leu | SNP | 97% | 62% (1011) |
| 1280201 | CDKN2A | c.237_238delCCinsTT | p.Arg80* | INDEL | 85% | 60% (1823) |
| 2420801 | CDKN2A | c.238C>T | p.Arg80* | SNP | 100% | 52% (1886) |
| 4320801 | CDKN2A | c.242C>T | p.Pro81Leu | SNP | 93% | 51% (1404) |
| 6420401 | CDKN2A | c.148C>T | p.Gln50* | SNP | 85% | 39% (868) |
| 6520601 | CDKN2A | c.181G>T | p.Glu61* | SNP | 96% | 91% (701) |
| 0870201 | CTNNB1 | c.95A>T | p.Asp32Val | SNP | 72% | 29% (1127) |
| 2510211 | CTNNB1 | c.110C>G | p.Ser37Cys | SNP | 50% | 25% (690) |
| 5400121 | CTNNB1 | c.98C>T | p.Ser33Phe | SNP | 50% | 21% (305) |
| 6100131 | CTNNB1 | c.110C>G | p.Ser37Cys | SNP | 45% | 35% (1069) |
| 2350301 | EGFR | c.2495G>A | p.Arg832His | SNP | 58% | 35% (2599) |
| 2510211 | EGFR | c.2126A>C | p.Glu709Ala | SNP | 80% | 66% (6589) |
| 2510211 | EGFR | c.2155G>A | p.Gly719Ser | SNP | 85% | 64% (6554) |
| 2510211 | EGFR | c.2369C>T | p.Thr790Met | SNP | 86% | 60% (3569) |
| 3800431 | EGFR | c.2281G>A | p.Asp761Asn | SNP | 44% | 28% (880) |
| 6770101 | EGFR | c.2236_2250delGAATTAAGAGAAGCA | p.Glu746_Ala750del | INDEL | 61% | 46% (1961) |
| 6770101 | EGFR | c.2369C>T | p.Thr790Met | SNP | 20% | 8% (560) |
| 6770102 | EGFR | c.2390G>C | p.Cys797Ser | SNP | 6% | 9% (2118) |
| 6770102 | EGFR | c.2236_2250delGAATTAAGAGAAGCA | p.Glu746_Ala750del | INDEL | 94% | 88% (9127) |
| 6770102 | EGFR | c.2369C>T | p.Thr790Met | SNP | 63% | 56% (13490) |
| 0920401 | ERBB2 | c.2329G>T | p.Val777Leu | SNP | 68% | 33% (1218) |
| 2350301 | ERBB2 | c.2317G>A | p.Val773Met | SNP | 50% | 37% (1653) |
| 6200511 | EZH2 | c.1937A>T | p.Tyr646Phe | SNP | 48% | 23% (279) |
| 6220401 | FBXW7 | c.1394G>A | p.Arg465His | SNP | 31% | 21% (1149) |
| 7930501 | FBXW7 | c.1513C>T | p.Arg505Cys | SNP | 26% | 33% (1150) |
| 2350301 | FGFR2 | c.1168G>A | p.Ala390Thr | SNP | 55% | covered, not called |
| 8600431 | FGFR2 | c.755C>G | p.Ser252Trp | SNP | 49% | 21% (458) |
| 5700111 | GNAS | c.602G>A | p.Arg201His | SNP | 56% | 37% (1416) |
| 5870201 | HRAS | c.37_38delGGinsAA | p.Gly13Asn | INDEL | 45% | 31% (602) |
| 0870201 | IDH1 | c.395G>A | p.Arg132His | SNP | 96% | 32% (863) |
| 1530701 | IDH1 | c.395G>A | p.Arg132His | SNP | 47% | 41% (521) |
| 6930501 | IDH1 | c.395G>A | p.Arg132His | SNP | 51% | 44% (856) |
| 0500121 | KRAS | c.38G>A | p.Gly13Asp | SNP | 100% | 68% (1860) |
| 0500831 | KRAS | c.35G>A | p.Gly12Asp | SNP | 58% | 40% (907) |
| 0700111 | KRAS | c.34G>A | p.Gly12Ser | SNP | 51% | 16% (227) |
| 1320601 | KRAS | c.34G>T | p.Gly12Cys | SNP | 58% | 32% (461) |
| 3220601 | KRAS | c.35G>A | p.Gly12Asp | SNP | 60% | 32% (544) |

| ID | Gene | coding | protein | type | WGS (tumor adjusted VAF) | Oncomine NGS panel VAF (coverage) |
|---|---|---|---|---|---|---|
| 3700111 | KRAS | c.35G>A | p.Gly12Asp | SNP | 57% | 30% (607) |
| 5700111 | KRAS | c.35G>A | p.Gly12Asp | SNP | 63% | 35% (618) |
| 6100131 | KRAS | c.34G>T | p.Gly12Cys | SNP | 48% | 32% (862) |
| 6930501 | KRAS | c.35G>A | p.Gly12Asp | SNP | 9% | 6% (338) |
| 7100131 | KRAS | c.356A>G | p.Asp119Gly | SNP | 33% | 25% (1477) |
| 7310211 | KRAS | c.35G>T | p.Gly12Val | SNP | 40% | 30% (929) |
| 7930501 | KRAS | c.35G>A | p.Gly12Asp | SNP | 58% | 54% (848) |
| 9620401 | KRAS | c.35G>T | p.Gly12Val | SNP | 72% | 22% (597) |
| 6420401 | MAP2K1 | c.371C>T | p.Pro124Leu | SNP | 35% | 11% (785) |
| 5870201 | MET | c.3065G>A | p.Arg1022Gln | SNP | 53% | 33% (3302) |
| 6200511 | MET | c.1130T>C | p.Ile377Thr | SNP | 43% | 31% (278) |
| 2420801 | NRAS | c.182A>G | p.Gln61Arg | SNP | 69% | 34% (512) |
| 3800431 | NRAS | c.181C>A | p.Gln61Lys | SNP | 41% | 24% (376) |
| 1320601 | PIK3CA | c.1624G>C | p.Glu542Gln | SNP | 49% | 15% (276) |
| 1530701 | PIK3CA | c.1636C>A | p.Gln546Lys | SNP | 19% | 13% (370) |
| 2420801 | PIK3CA | c.1633G>A | p.Glu545Lys | SNP | 52% | 30% (1094) |
| 3220601 | PIK3CA | c.1624G>A | p.Glu542Lys | SNP | 49% | 15% (460) |
| 3800431 | PIK3CA | c.1624G>A | p.Glu542Lys | SNP | 57% | 22% (540) |
| 5400121 | PIK3CA | c.1633G>A | p.Glu545Lys | SNP | 54% | 16% (314) |
| 6100131 | PIK3CA | c.1624G>A | p.Glu542Lys | SNP | 50% | 25% (685) |
| 6930501 | PIK3CA | c.1624G>A | p.Glu542Lys | SNP | 10% | 8% (350) |
| 7310211 | PIK3CA | c.1633G>A | p.Glu545Lys | SNP | 59% | 29% (1018) |
| 8250301 | PIK3CA | c.1624G>A | p.Glu542Lys | SNP | 37% | 25% (679) |
| 8600431 | PIK3CA | c.3140A>G | p.His1047Arg | SNP | 84% | 71% (11177) |
| 2350301 | POLE | c.1270C>G | p.Leu424Val | SNP | 62% | 34% (600) |
| 0500121 | PTEN | c.740_741insA | p.Pro248fs | INDEL | 92% | 42% (1647) |
| 2510211 | PTEN | c.406T>C | p.Cys136Arg | SNP | 100% | 21% (405) |
| 6370201 | PTEN | c.389G>A | p.Arg130Gln | SNP | 100% | 55% (930) |
| 6420401 | PTEN | c.980A>G | p.Lys327Arg | SNP | not reported | 10% (155) |
| 9180201 | PTEN | c.80A>C | p.Tyr27Ser | SNP | 12% | covered, not reported |
| 2420801 | RAF1 | c.770C>T | p.Ser257Leu | SNP | 44% | 33% (1191) |
| 6930501 | RAF1 | c.779C>T | p.Thr260Ile | SNP | 16% | 13% (397) |
| 0500121 | SMAD4 | c.1082G>A | p.Arg361His | SNP | 100% | 55% (1134) |
| 1280201 | SMAD4 | c.325C>G | p.Leu109Val | SNP | 29% | 24% (1265) |
| 3700111 | SMAD4 | c.1081C>T | p.Arg361Cys | SNP | 100% | 34% (773) |
| 6100131 | SMAD4 | c.1082G>A | p.Arg361His | SNP | 46% | 35% (1050) |
| 6220401 | SMAD4 | c.353C>T | p.Ala118Val | SNP | 93% | 42% (1288) |
| 7280101 | SMAD4 | c.1082G>A | p.Arg361His | SNP | 100% | 9% (215) |
| 0500121 | TP53 | c.743G>A | p.Arg248Gln | SNP | 76% | 55% (2092) |
| 0500831 | TP53 | c.743G>A | p.Arg248Gln | SNP | 78% | 49% (2683) |
| 0500831 | TP53 | c.843C>A | p.Asp281Glu | SNP | 28% | 25% (398) |
| 0700111 | TP53 | c.916C>T | p.Arg306* | SNP | 100% | 21% (400) |
| 1320601 | TP53 | c.404G>A | p.Cys135Tyr | SNP | 97% | 27% (536) |
| 1530701 | TP53 | c.404G>A | p.Cys135Tyr | SNP | 100% | 77% (1263) |
| 2420801 | TP53 | c.743G>A | p.Arg248Gln | SNP | 100% | 55% (1698) |
| 3220601 | TP53 | c.742C>T | p.Arg248Trp | SNP | 100% | 27% (820) |
| 4320801 | TP53 | c.1023_1024delCCinsTT | p.Arg342* | INDEL | 99% | 47% (2048) |
| 4410311 | TP53 | c.422G>A | p.Cys141Tyr | SNP | 100% | 66% (1767) |
| 4980101 | TP53 | c.713G>T | p.Cys238Phe | SNP | 93% | 32% (1175) |
| 5400121 | TP53 | c.817C>T | p.Arg273Cys | SNP | 75% | 44% (584) |
| 5870201 | TP53 | c.1009C>G | p.Arg337Gly | SNP | 91% | 66% (2232) |
| 6770101 | TP53 | c.527G>T | p.Cys176Phe | SNP | 100% | 41% (1860) |
| 6770101 | TP53 | c.527G>T | p.Cys176Phe | SNP | 100% | 69% (2492) |
| 6930501 | TP53 | c.817C>T | p.Arg273Cys | SNP | 99% | 91% (5809) |
| 7280101 | TP53 | c.817C>T | p.Arg273Cys | SNP | 89% | 17% (816) |
| 7310211 | TP53 | c.853G>A | p.Glu285Lys | SNP | 74% | 37% (791) |
| 7930501 | TP53 | c.524G>A | p.Arg175His | SNP | 100% | 73% (1778) |
| 9110211 | TP53 | c.818G>A | p.Arg273His | SNP | 91% | 55% (1997) |
| 9200721 | TP53 | c.824G>T | p.Cys275Phe | SNP | 83% | 45% (866) |
| 9620401 | TP53 | c.818G>A | p.Arg273His | SNP | 100% | 31% (1299) |
