## Supplementary Table 6 for "Clinical validation of Whole Genome Sequencing for cancer diagnostics"

| sample ID | Tumor type | CDKN2A/p16 IHC | CDKN2A WGS |
|---|---|---|---|
| 6530103 | Sarcoma | negative (minor) | 0 copies |
| 6360103 | Cancer of unknown primary | negative | 0 copies |
| 2140103 | Bladder cancer | negative | 0 copies |
| 6120103 | Ovarian cancer | negative | 0 copies |
| 9110103 | Cancer of unknown primary | negative | 0 copies |
| 2110103 | Vaginal cancer | negative | 0 copies (partial gene) |
| 4060103 | Cancer of unknown primary | negative | 0 copies (partial gene) |
| 7990103 | Cancer of unknown primary | negative | 0 copies |
| 0480103 | Lung cancer | negative | 0 copies |
| 6030103 | Lung cancer | negative | 0 copies |
| 9330103 | Breast Cancer | negative | 0 copies |
| 7330103 | Cancer of unknown primary | negative | 0 copies (partial gene) |
| 4330103 | Lung cancer | negative | 0 copies |
| 3330103 | Lung cancer | negative | 0 copies |
| 1301103 | Bladder cancer | negative | 0 copies |
| 2580103 | Lung cancer | negative | 0 copies |
| 8380103 | Lung cancer | negative | 0 copies |
| 6560103 | Lung cancer | negative | 0 copies |
| 6130103 | Lung cancer | negative | 0 copies |
| 3130103 | Lung cancer | negative | 0 copies |
| 4201103 | Lung cancer | negative | 0 copies |
| 1380103 | Lung cancer | negative | 0 copies |
| 7000103 | Cervical cancer | positive | 2 copies (wildtype) |
| 5710103 | Lung cancer | positive (weaker) | 2 copies (wildtype) |
| 4730103 | Sarcoma | positive | 1 copy (wildtype) |
| 9950103 | Anal cancer | positive | 4 copies (wildtype) |
| 0750103 | Cervical cancer | positive (weaker) | 4 copies (wildtype) |
| 4160103 | Cancer of unknown primary | positive (weaker) | 4 copies (wildtype) |
| 5950103 | Anal cancer | positive | 3 copies (wildtype) |
| 0450103 | Thymoma and Thymic Cancer | positive (weaker) | 2 copies (wildtype) |
| 2280103 | Sarcoma | positive (weaker) | 1 copy (wildtype) |
| 2780103 | Anal cancer | positive | 2 copies (wildtype) |
| 3190103 | Sarcoma | positive | 4 copies (wildtype) |
| 1990103 | Ovarian cancer | positive (weaker) | 2 copies (wildtype) |
| 0001103 | Bladder cancer | positive | 4 copies (wildtype) |
| 4280103 | Penile cancer | positive (weaker) | 4 copies (wildtype) |
| 7201103 | Head and Neck cancer | positive | 2 copies (wildtype) |
| 0701103 | Cervical cancer | positive (weaker) | 4 copies (wildtype) |
| 6901103 | Cancer of unknown primary | positive (weaker) | 1 copy (wildtype) |
